# Electrophysiological Studies of Reception of Facial Communication in Autism Spectrum Disorder and Schizophrenia

**DOI:** 10.1101/19013003

**Authors:** Emily J. Levy, Jennifer Foss-Feig, Emily L. Isenstein, Vinod Srihari, Alan Anticevic, Adam J. Naples, James C. McPartland

## Abstract

Autism spectrum disorder (ASD) and schizophrenia spectrum disorders (SZ) are both characterized by difficulty with social cognition. Likewise, social brain activity is atypical in both disorders and indicates atypical reception of facial communication – a key area in the Research Domain Criteria framework for identifying common biological underpinnings of psychiatric disorders. To identify areas of overlap and dissociation between ASD and SZ, this paper reviews studies of electrophysiological (EEG) response to facial stimuli across ASD and SZ populations. We focus on findings regarding amplitude and latency of four brain responses implicated in social perception: P100, N170, N250, and P300. There were many inconsistent findings in both the ASD and SZ literatures; however, replication across studies was strongest for delayed N170 latency in ASD and attenuated N170 amplitude in SZ. EEG responses corresponded with clinical symptoms in multiple samples. These results highlight the challenges associated with replicating research findings in heterogeneous clinical populations, as well as the need for transdiagnostic research and for designing studies to examine relationships among continuous quantifications of behavior and neural activity across neurodevelopmental disorders.

## INTRODUCTION

Autism spectrum disorder (ASD) and schizophrenia spectrum disorders (SZ) are highly prevalent neurodevelopmental disorders (ND) with considerable public health impact, costing the United States a combined $423 billion per year (Cloutier et al. 2016). Though these disorders are classified in two separate diagnostic taxonomies, social deficits are diagnostic hallmarks of both, and diagnostic confusion is common. Social dysfunction is a core symptom of both ASD and SZ, evident in decreased social motivation, reduced social reward, impaired mentalizing, and decreased likelihood of social engagement (Dawson et al. 2005; Dowd and Barch 2010; Schultz 2005). Moreover, parallel lines of research indicate commonalities in affected genetic pathways and neural processes, suggesting shared neuropathology (Cristino et al. 2014; Mitchell 2011). These findings are consistent with the hypothesis that common mechanisms contribute to social communication deficits across behaviorally defined categories of ND. Despite these commonalities, few studies have directly compared distinct diagnostic groups or conceptualized these diagnostic classes as heterogeneous manifestations of shared dysfunction in overlapping neural substrates. For this reason, little is known about whether atypical social perception across NDs reflects common or distinct neural underpinnings. In this review, we focus on literature related to face and emotion processing across ASD and SZ, highlighting what is known, where the disorders converge and diverge, and what work remains to be done to understand social processes related to reception of facial communication across disorders.

Decreased attention to human faces is evident throughout the lifespan in ASD (Maestro et al. 2002; Osterling and Dawson 1994; Dawson et al. 2005). Though SZ is not usually diagnosed until adolescence or early adulthood, infants later diagnosed with psychosis also show decreased attention to faces (Massie 1978). Eye-tracking studies of perception of facial stimuli demonstrate atypical face scanning in individuals with ASD (Pelphrey et al. 2002; A. Klin et al. 2002; Joseph and Tanaka 2003) and SZ (Phillips and David 1997; Gordon et al. 1992), with both clinical groups spending less time fixating on the eyes and more time fixating on other parts of the face. Individuals with ASD (Ami Klin et al. 1999; Hobson 1986; Baron-Cohen et al. 2001; Law Smith et al. 2010) and SZ (Bellack et al. 1996; Mueser et al. 1996; Morrison et al. 1988) also show deficits in accurately identifying emotions, sorting emotional faces, and matching faces based on emotion. Atypical sensitivity to emotional expressions is a stable feature of multiple NDs (Addington and Addington 1998; Gaebel and Wolwer 1992) and has been shown to associate with chronicity (Kington et al. 2000), severity, and type of active symptomatology (Davis and Gibson 2000; Lewis and Garver 1995). Difficulties in emotion recognition and discrimination in SZ and ASD are associated with specific disruption in social process systems, such as increased social withdrawal and impaired social cognition.

Face and emotion processing rely upon a network of specialized brain regions. Neuroimaging research has clarified the role of specific brain areas, including the fusiform gyrus, superior temporal sulcus, and amygdala, in face perception (Kesler-West et al. 2001; Kanwisher 2001). EEG studies reveal a specific time course for related but distinct stages of face processing in this network (T. Wong et al. 2009; Luo et al. 2009). This sequence of event-related potentials (ERPs) is distinguished by scalp topography and chronology and shows sensitivity across perceptual processes, from detecting faces (S. Bentin et al. 1996) to interpreting differences in facial identity, direction of gaze, and displays of emotion (Conty et al. 2007; Gosling and Eimer 2011; M. Eimer and Holmes 2002). These ERPs provide temporally precise indices of function across the regions involved in face perception, or, as conceptualized in the NIMH’s Research Domain Criteria (RDoC) framework, the subdomain *reception of facial communication* within the broader category of *social process systems* (Kozak and Cuthbert 2016). Prior research has demonstrated the relevance of a subset of these components across neurological and psychiatric disorders (Feuerriegel et al. 2015), and here we focus on their manifestation in ASD and SZ, reviewing the broad, temporally sequential set of ERP components associated with face processing (P100, N170, N250, and P300).

Measured over the occipital cortex as a positive deflection approximately 100ms after the onset of a visual stimulus, the P100 reflects low-level processing in the visual cortex (Mangun 1995; V1; A. P. F. Key et al. 2005) that is modulated by age and may be affected by the social nature of stimuli. In typical development (TD), faces elicit a larger (Herrmann et al. 2005; Roxane J Itier and Taylor 2004c) and faster (Kuefner et al. 2009; Taylor et al. 2001) P100 than objects, and in a number of studies, inverted faces elicit a larger (Roxane J Itier and Taylor 2004c, 2004a) but slower (Roxane J Itier and Taylor 2004c; Taylor et al. 2001) P100 than upright faces. P100 amplitude can be modulated by emotion (Magali Batty and Taylor 2003).

The N170 is a negative-going component, recorded over occipito-temporal scalp approximately 170ms after viewing a face, that indexes the earliest stages of face processing (i.e., structural encoding). Neural generators of the N170 have been localized to occipito-temporal sites, including the fusiform gyrus (B. Rossion et al. 2003; Sadeh et al. 2010; R. J. Itier and Taylor 2002), superior temporal sulcus (Roxane J Itier and Taylor 2004d; Yovel et al. 2008), lingual gyrus (Shibata et al. 2002), posterior inferotemporal gyrus (Shibata et al. 2002; Schweinberger et al. 2002), and inferior occipital gyrus (Jacques et al. 2018). Like the P100, in TD the N170 exhibits enhanced amplitude (more negative) and decreased latency to faces relative to objects (Roxane J Itier and Taylor 2004c; S. Bentin et al. 1996) and larger amplitude but slower latency to inverted than to upright faces (Martin Eimer 2000; Roxane J Itier and Taylor 2004b, 2004d, 2004c; Bruno Rossion et al. 2000). For the N170, this “face inversion effect” is not found for upright vs. inverted objects, consistent with the interpretation that the N170 indexes face-selective processing (Martin Eimer 2000; Roxane J Itier and Taylor 2004c). N170 latency tends to be faster (Blau et al. 2007) with greater amplitude (Roxane J Itier and Taylor 2004b, 2004c; B. Rossion et al. 2003) over the right hemisphere than the left. Finally, N170 also may reflect an early indicator of emotion processing. Happy faces elicit faster N170 latencies than faces displaying negative affect (Magali Batty and Taylor 2003), and fearful faces elicit more negative N170 amplitude than neutral (Magali Batty and Taylor 2003; Blau et al. 2007) and happy (C. A. Brenner et al. 2014) faces.

The N250 is a negative-going component occurring approximately 250ms post-stimulus (Schweinberger et al. 2002). Though less well characterized than the P100 and N170 components and rarely studied in typically developing populations, the N250 demonstrates increased amplitude in response to emotional expressiveness relative to neutral faces (Balconi and Pozzoli 2008; L. Carretié et al. 2001; Labuschagne et al. 2010; Sato et al. 2001; Streit et al. 2001). As such, the N250 is considered a marker of higher-order face processing, such as affect decoding and emotion processing, and is presumed to reflect the modulatory influence of subcortical structures, including the amygdala (Streit et al. 1999). Research on face learning and repetition suggests the N250 is modulated by the identity of a face; familiar or repeated faces evoke a more negative N250 than unfamiliar or novel faces (Gosling and Eimer 2011; Kaufmann et al. 2009; Tanaka et al. 2006). N250 latency delays to inverted faces have been observed in a face repetition task (Roxane J Itier and Taylor 2004b), and, as with the N170, N250 has been shown to be right lateralized (Kaufmann et al. 2009).

The P300 is a large-amplitude positive component measured at central locations approximately 300ms after stimulus onset. It is associated with detecting novel and significant events (T. Picton 1995; Polich et al. 1995; T. W. Picton 1992; Ferrari et al. 2010), as well as with context updating (Donchin 2008; Donchin and Coles 1998, 1988). In the context of face perception, the P300 reflects processing of self-relevant stimuli, such as one’s own face (Onitsuka et al. 2001; Ninomiya et al. 1998). Emotional stimuli elicit a greater P300 amplitude than neutral stimuli (V. S. Johnston et al. 1986), and salient or attended stimuli elicit a greater P300 response than passively viewed stimuli (L Carretié et al. 1997). P300 amplitude is larger to upright than to inverted faces and may be modified by emotional valence (Kestenbaum and Nelson 1992), though the evidence is mixed (Conroy and Polich 2007).

In this review, we highlight findings related to the amplitude and latency of these four ERP components, as elicited in response to face and emotion stimuli in individuals with ASD and SZ. This work expands on prior reviews focused solely on the N170 in ASD (Kang et al. 2018) and other disorders (Feuerriegel et al. 2015) by reviewing points of convergence and divergence for additional neural components associated with face processing transdiagnotically.

## METHODS

### Search Criteria

Scopus, Google Scholar, and PubMed were queried using the following search criteria: (ASD OR autis* OR “pervasive developmental disorder” OR PDD-NOS OR Asperger* OR SZ OR schizo*) AND (ERP OR EEG OR electrophys* OR event-related potential OR P100 OR N170 OR N250 OR P300) AND (face OR facial OR emotion*). Reference lists, review articles, and meta-analyses were scanned to identify additional peer-review articles. Searches reflect updated content through October 23, 2017.

### Inclusion Criteria: Methods and Components

These searches yielded 69 eligible studies (36 ASD, 33 SZ); all included articles were peer-reviewed. Eligible studies were required to investigate at least one of the four key ERP components most commonly investigated in neuroscientific studies of face processing in typical and atypical development: P100, N170, N250, and/or P300. Studies that did not include at least one of these components were excluded (e.g., McMahon and Henderson 2015). Included studies were required to use images of faces as stimuli. Studies in which more than one face or object was simultaneously presented were excluded (e.g., Maher et al. 2016), as were studies in which all visual stimuli were combined with auditory stimuli (Müller et al. 2012). Any study that included faces stimuli was designated as a ‘face’ study, and any study that included face stimuli and contrasted response to two or more emotions was designated as an ‘emotion’ study. All ‘emotion’ studies that examined main effects of group collapsed across different emotions were also included as ‘face’ studies.

### Inclusion Criteria: Subjects

Details about population demographics, subject characterization, study methodology, and stimuli are presented in Table 1. Studies were required to examine individuals diagnosed with either ASD or SZ, and those studying only high-risk individuals or relatives were excluded (Wolwer et al. 2012). No studies included both diagnostic groups. Studies were required to report their method for confirming patients’ diagnostic status. In half of the studies of ASD (18 of 36), diagnosis was confirmed with a gold-standard clinical assessment, the Autism Diagnostic Observation Schedule (ADOS; Lord et al. 2012). For the remaining ASD studies, participants entered the study with a prior diagnosis according to DSM-IV or DSM-5 criteria. In 32 of 33 SZ studies, clinicians confirmed diagnosis based on DSM-IV criteria. The single study that did not specify the use of DSM criteria for schizophrenia completed a Schedule for Schizophrenia and Affective Disorders (Spitzer and Endicott 1979). Study samples targeting individuals under three years of age were excluded, given the developmental emergence of some of the ERP components at this point (Roxane J Itier and Taylor 2004b). Studies were otherwise included regardless of target age group; however, all SZ studies targeted adults or late adolescents, whereas two-thirds of ASD studies exclusively included children. Studies that included both a child and an adult group analyzed separately were treated as two separate studies. Studies without a typically developing (TD) comparison group were excluded. Studies examining additional clinical groups not targeted by this review (Tye et al. 2014; Tye et al. 2013) were included, but we focused only on results regarding ASD and SZ.

**Table 1.**
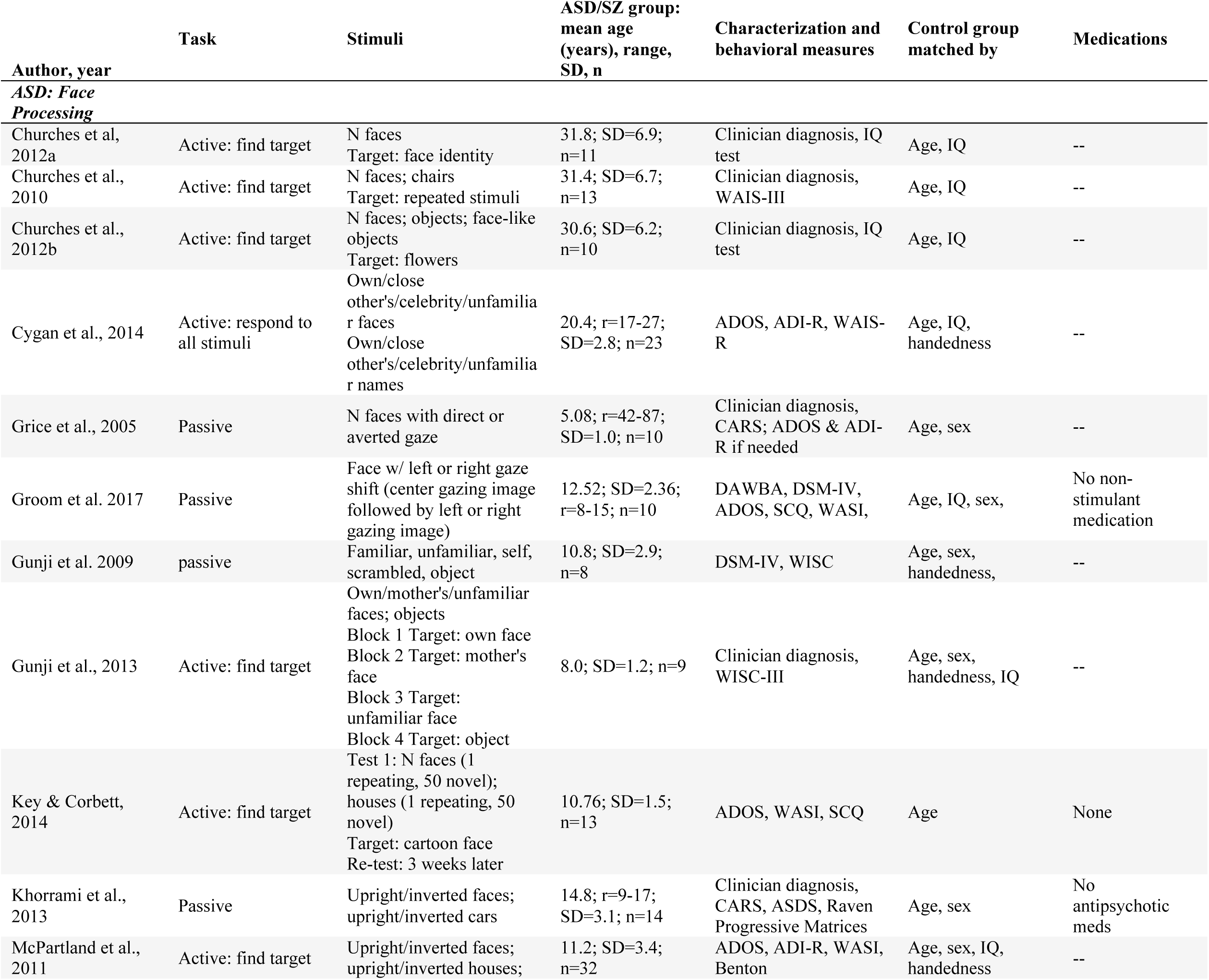
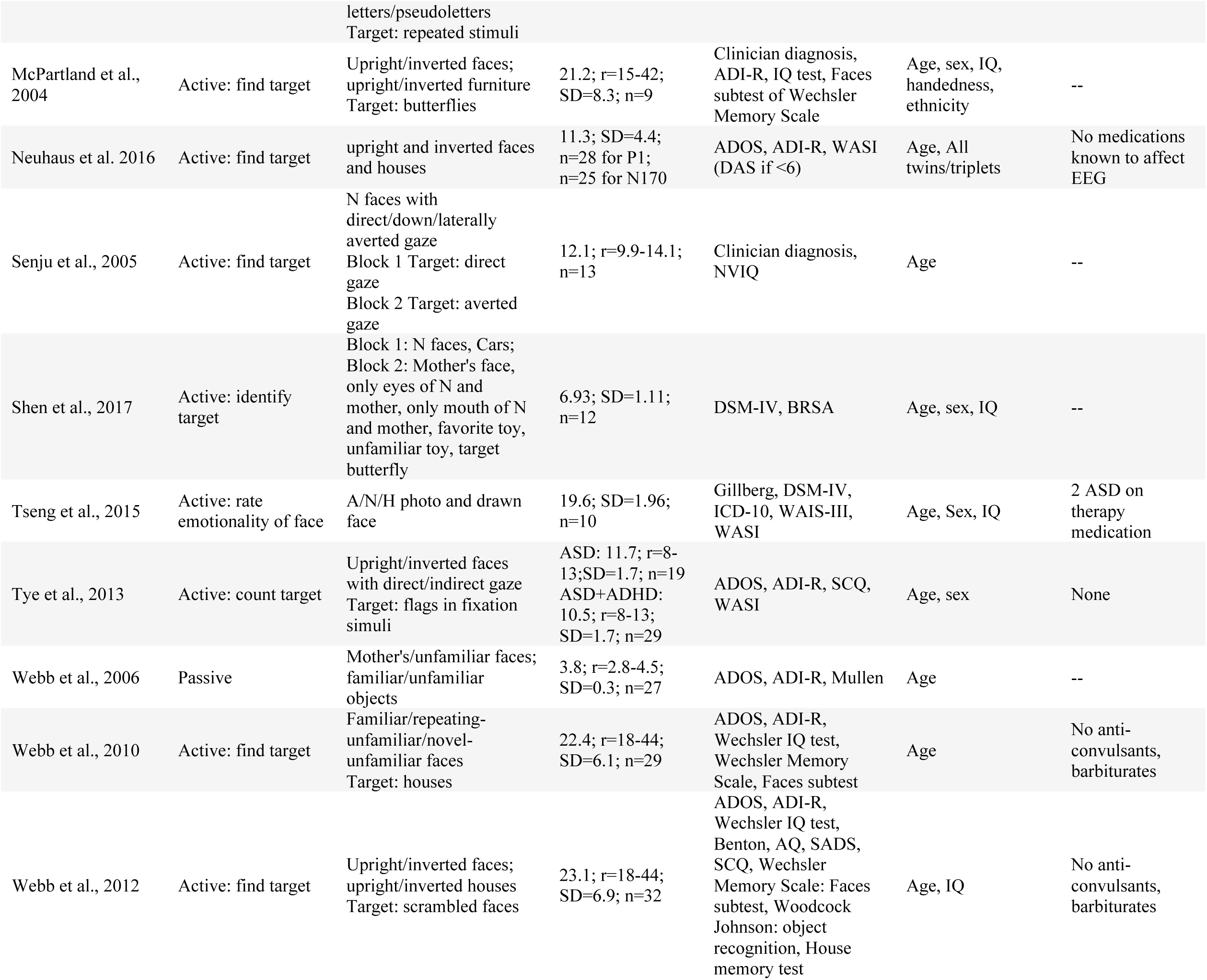
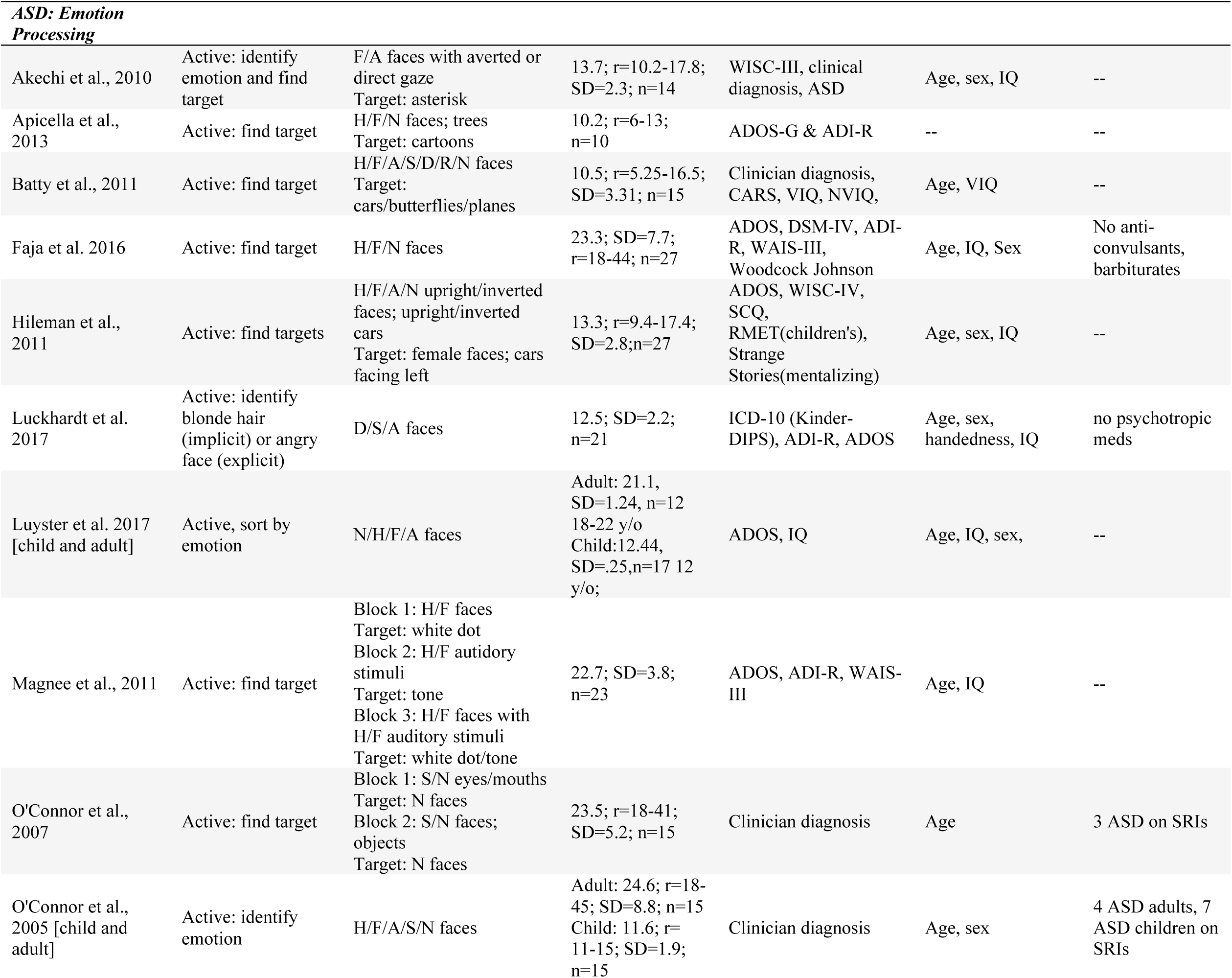
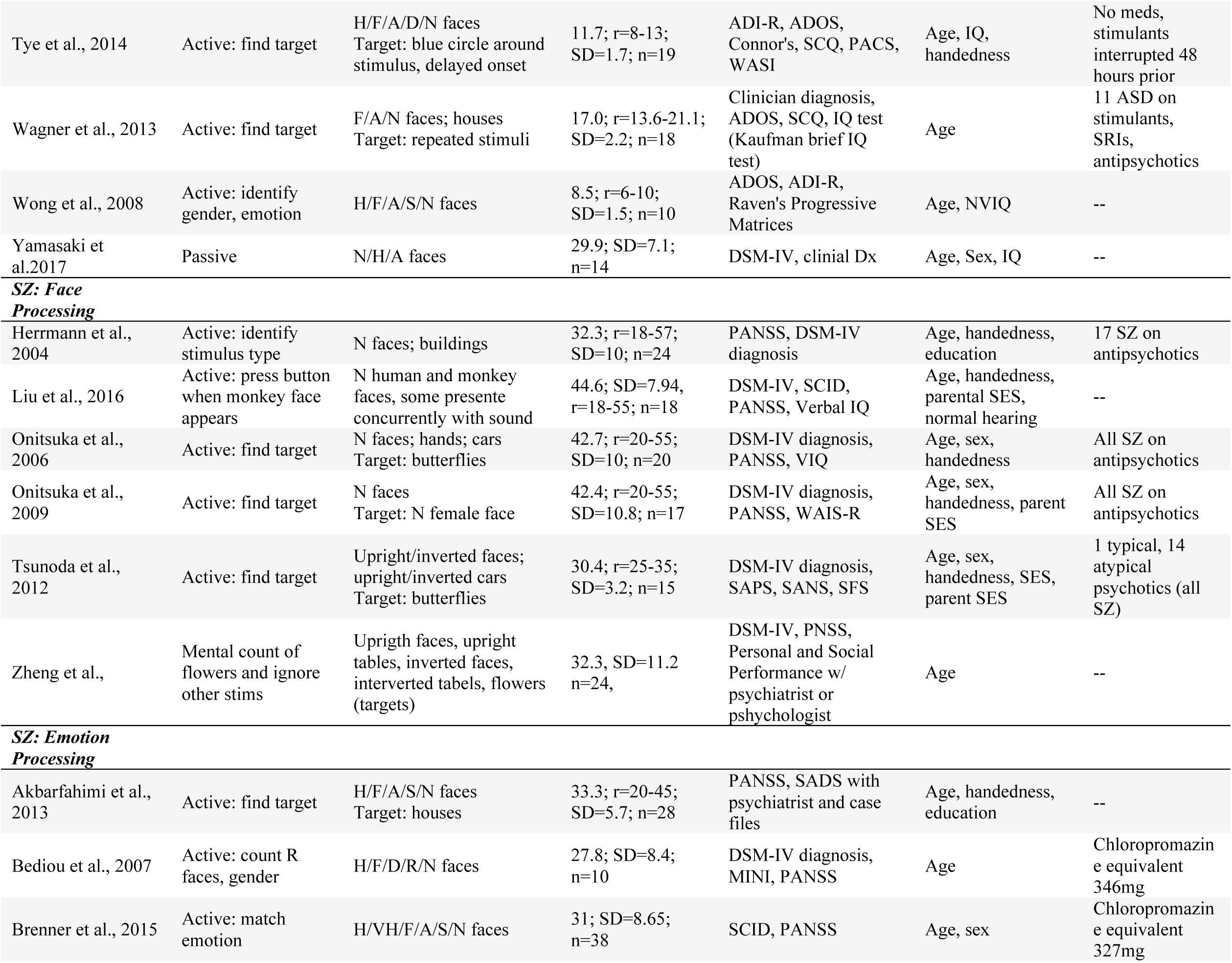
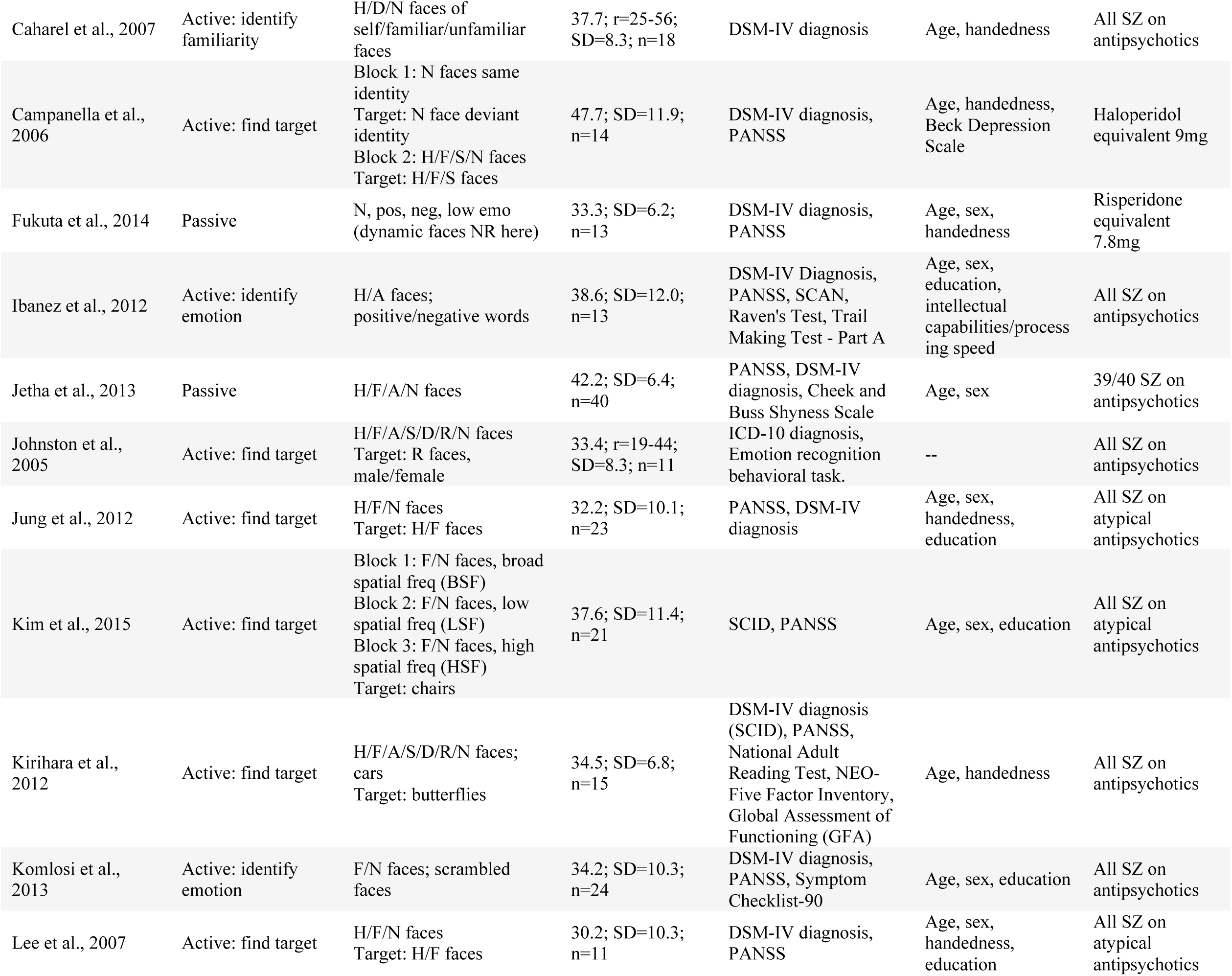
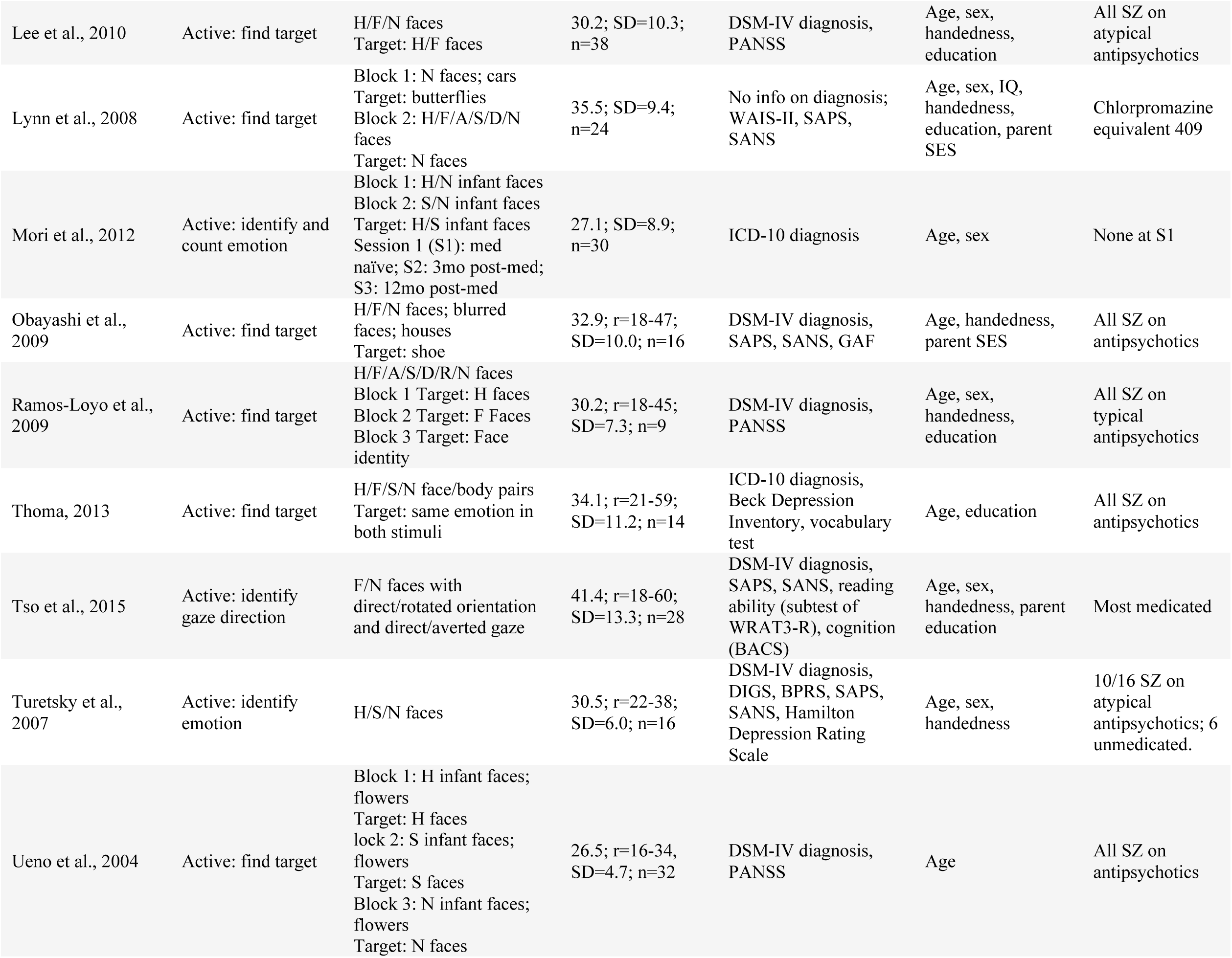
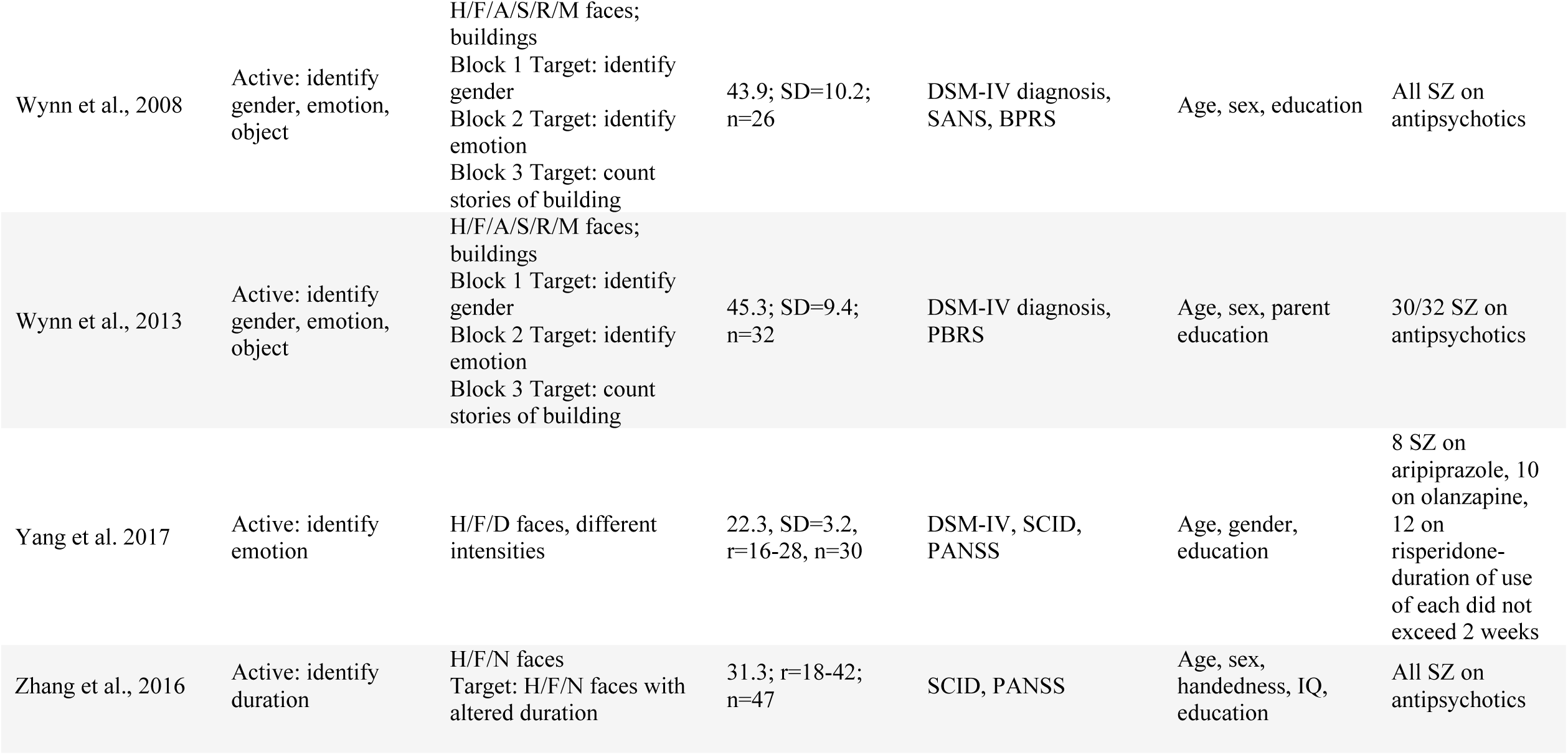
Participant characterization and ERP methods for all studies reviewed. Angry (A), Attention Deficit-Hyperactivity Disorder group (ADHD), amplitude (amp), Autism Spectrum Disorder group (ASD), disgusted (D), emotion (emo), fear (F), face-only effect (fo; study does not include non-face stimuli), happy (H), latency (lat), ashamed (M), neutral (N), negative (neg), not face-specific effect (nfs; effect across face and non-face stimuli), object (obj), positive (pos), surprised (R), sad (S), schizophrenia group (SZ), typically developing group (TD). Note: “Targets” were not included in analyses unless repeated in cell.

## RESULTS

Details on all study results described below can be found in Table 2. A graphical overview of findings can be found in Figure 1.

**Table 2.**
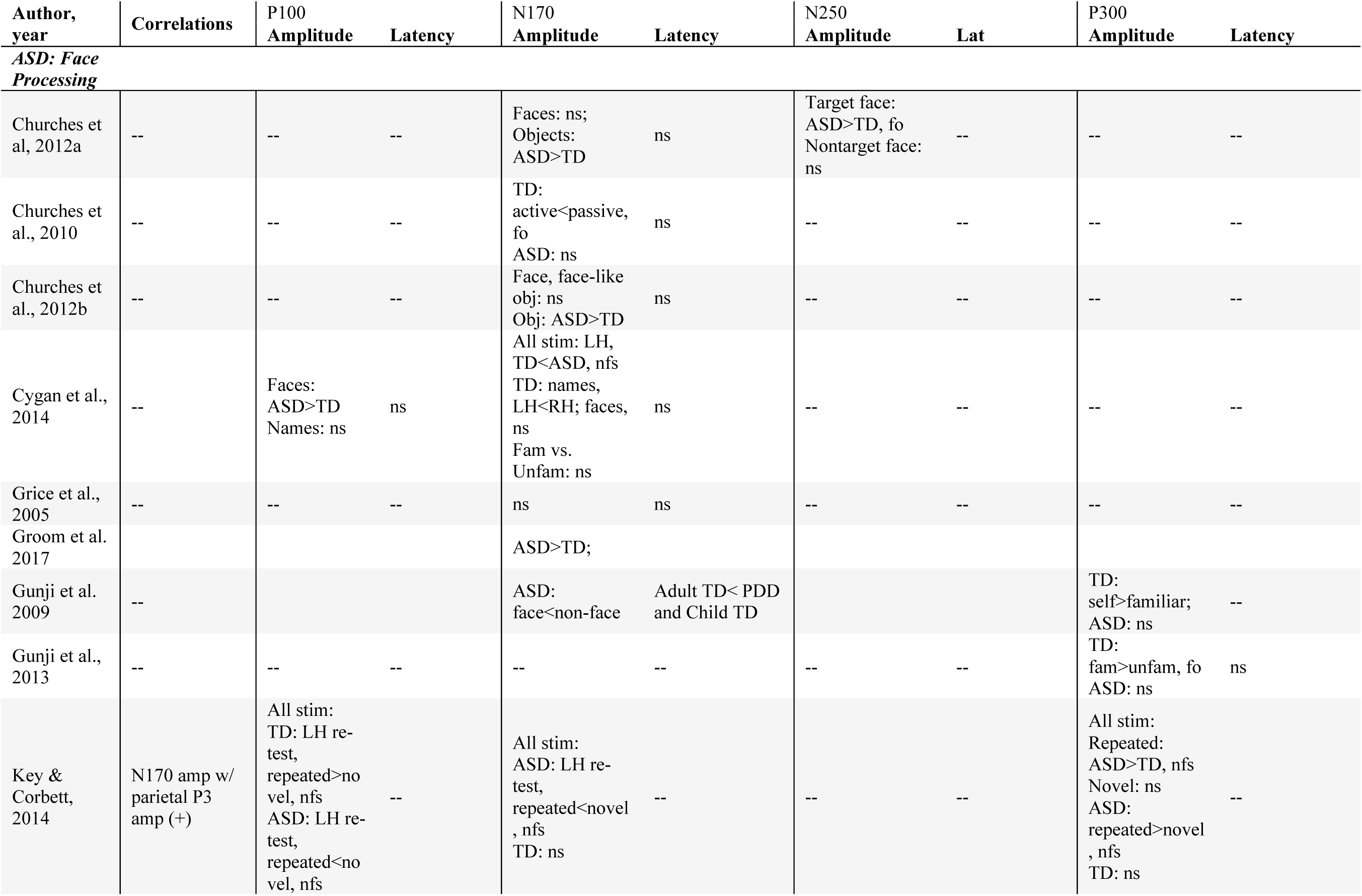
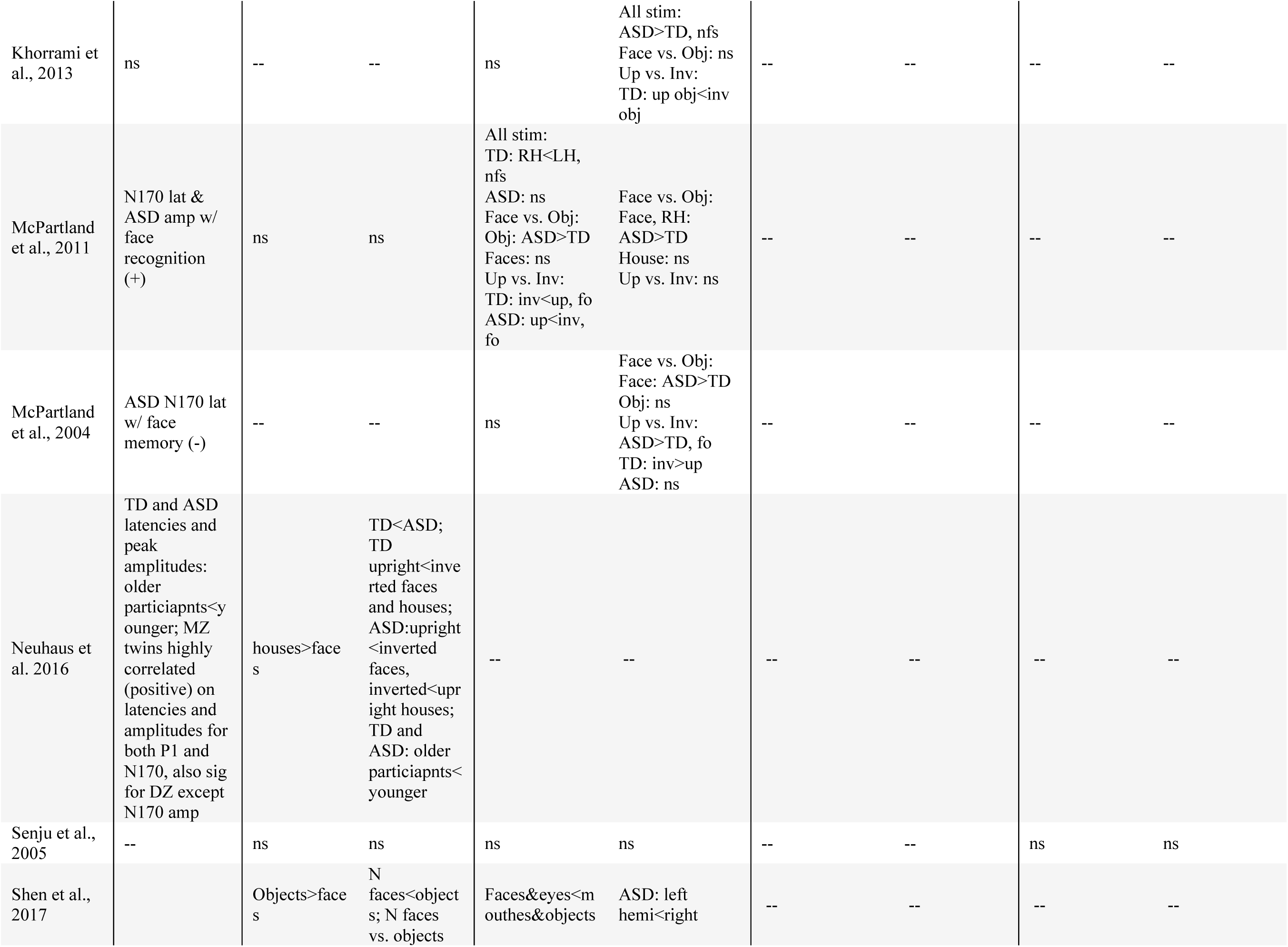
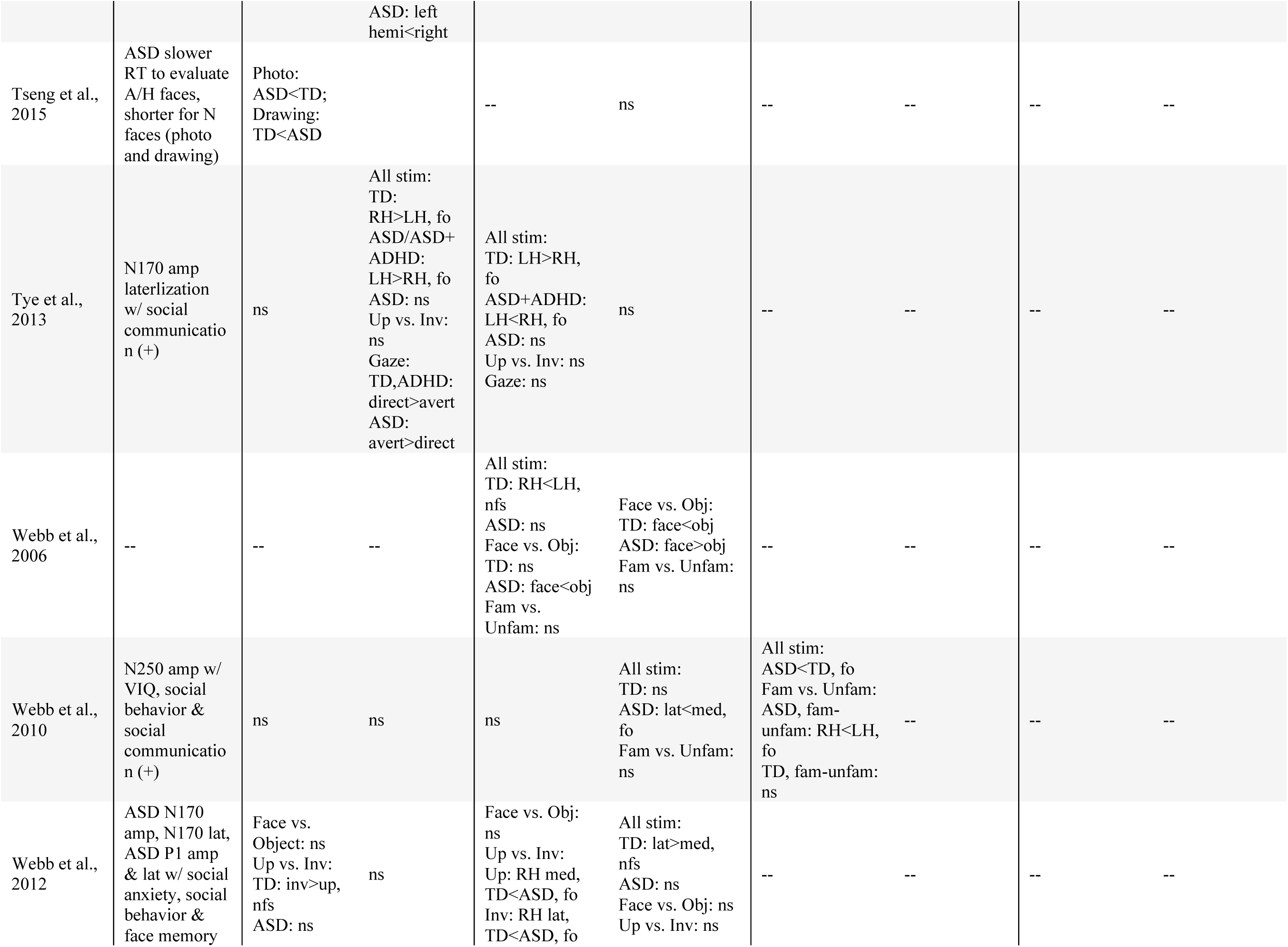
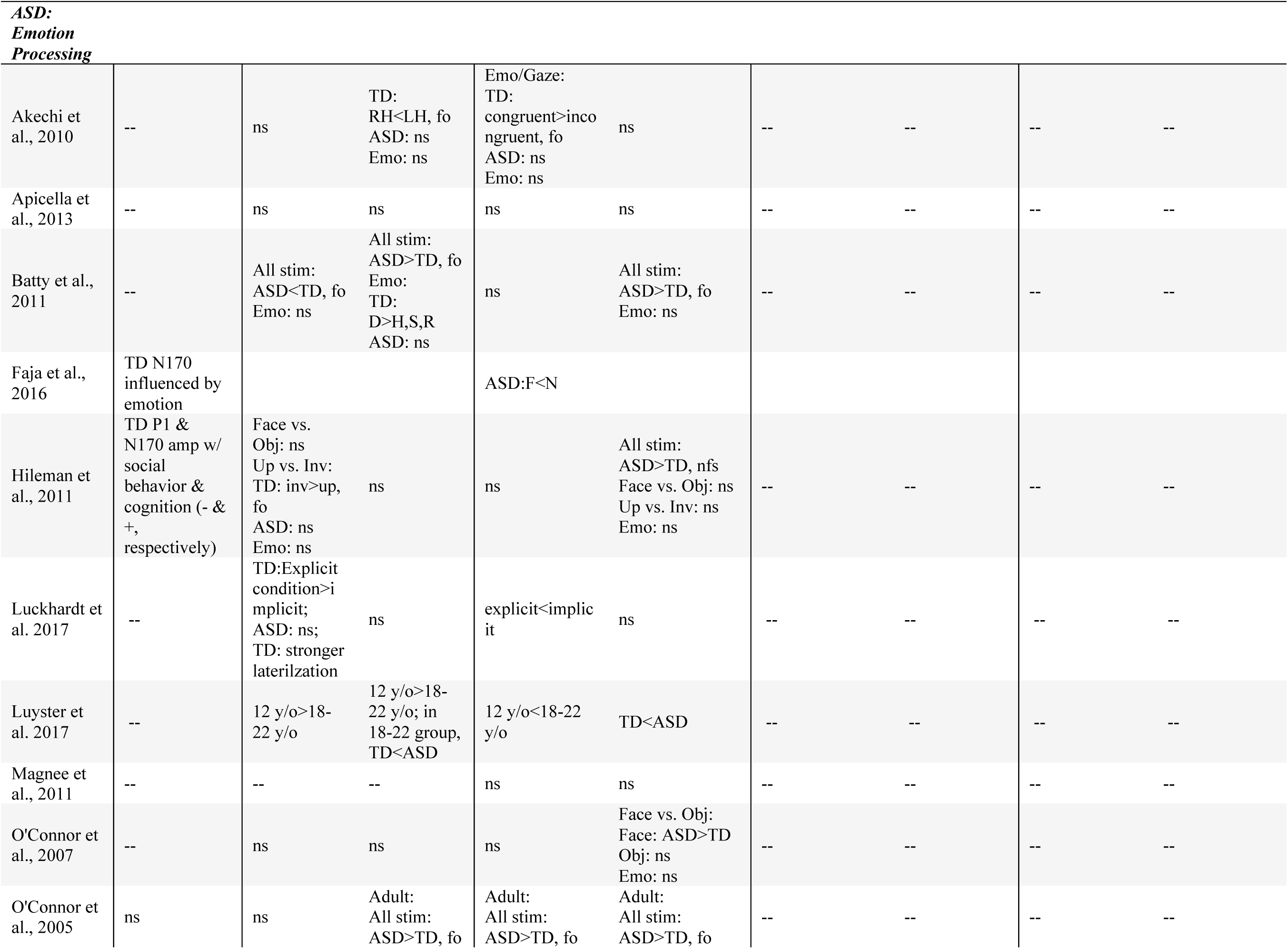
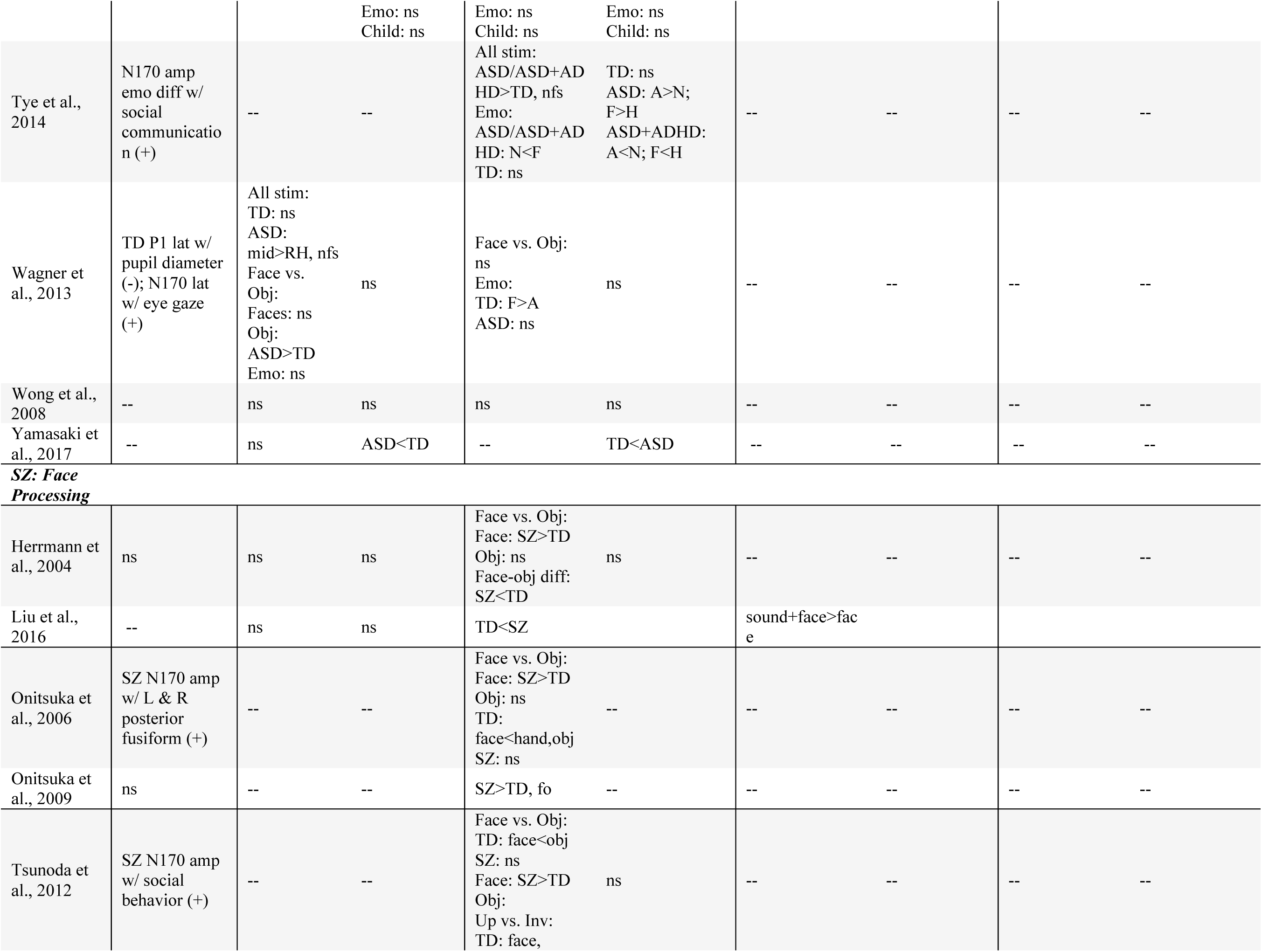
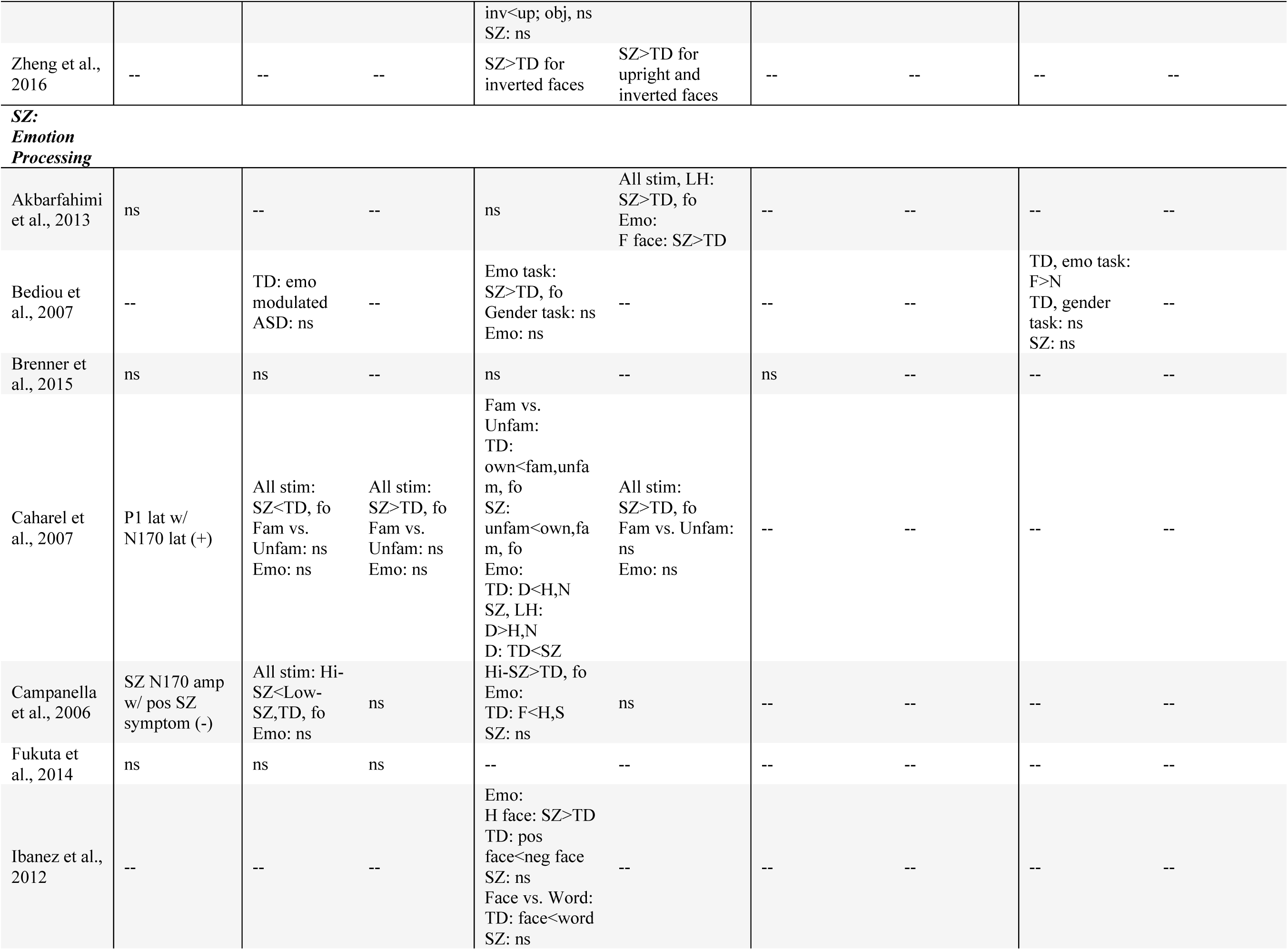
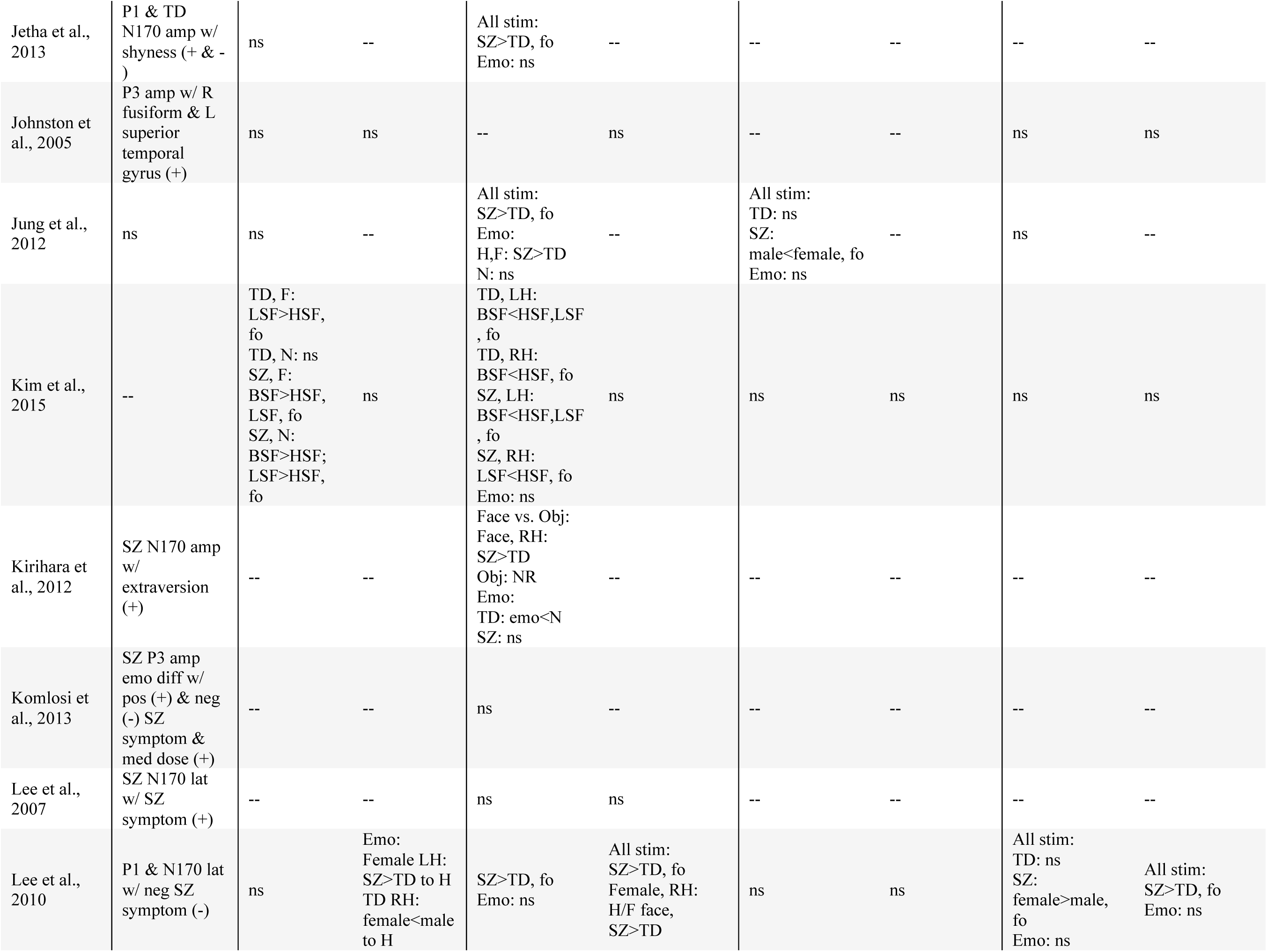
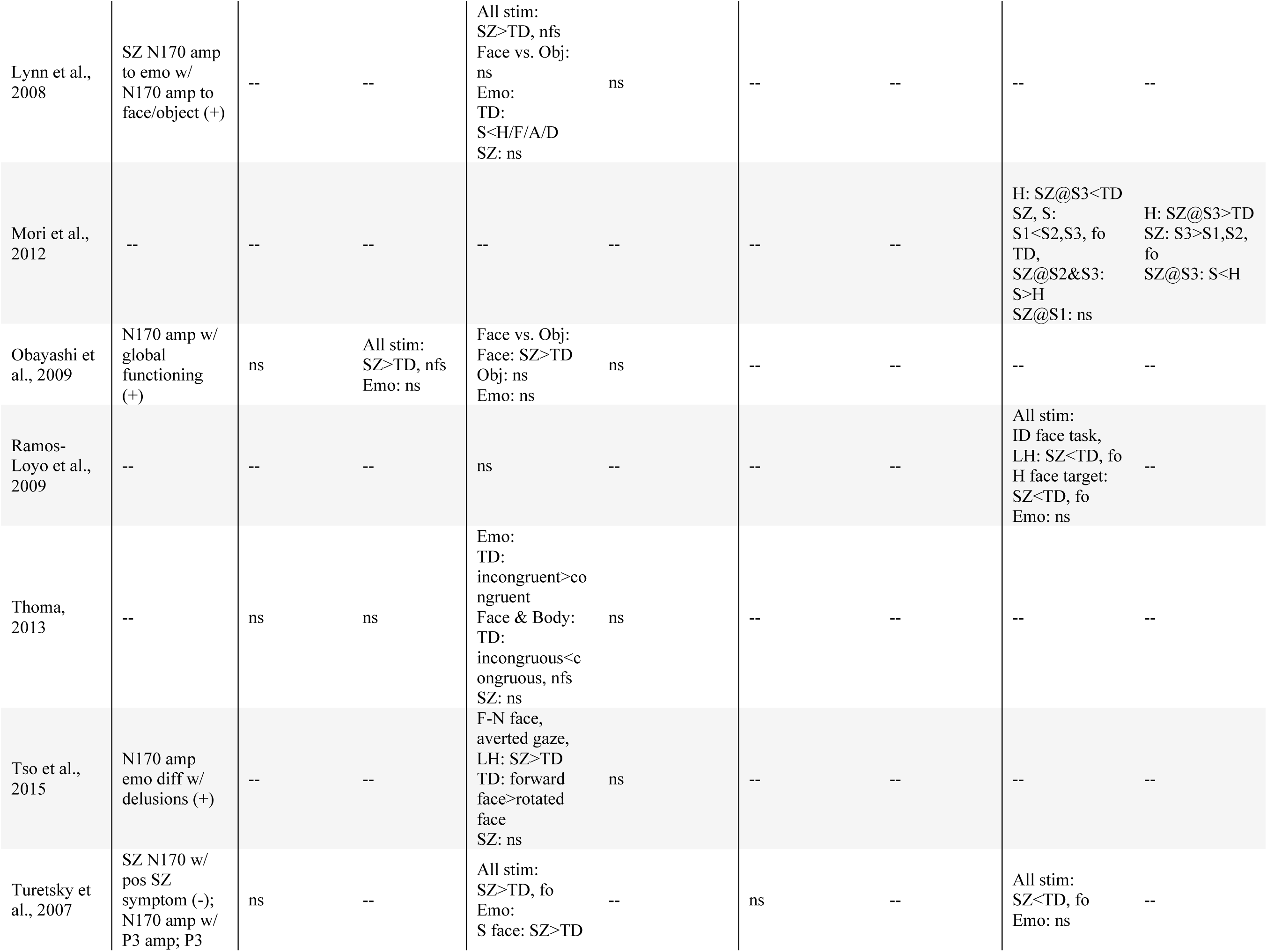
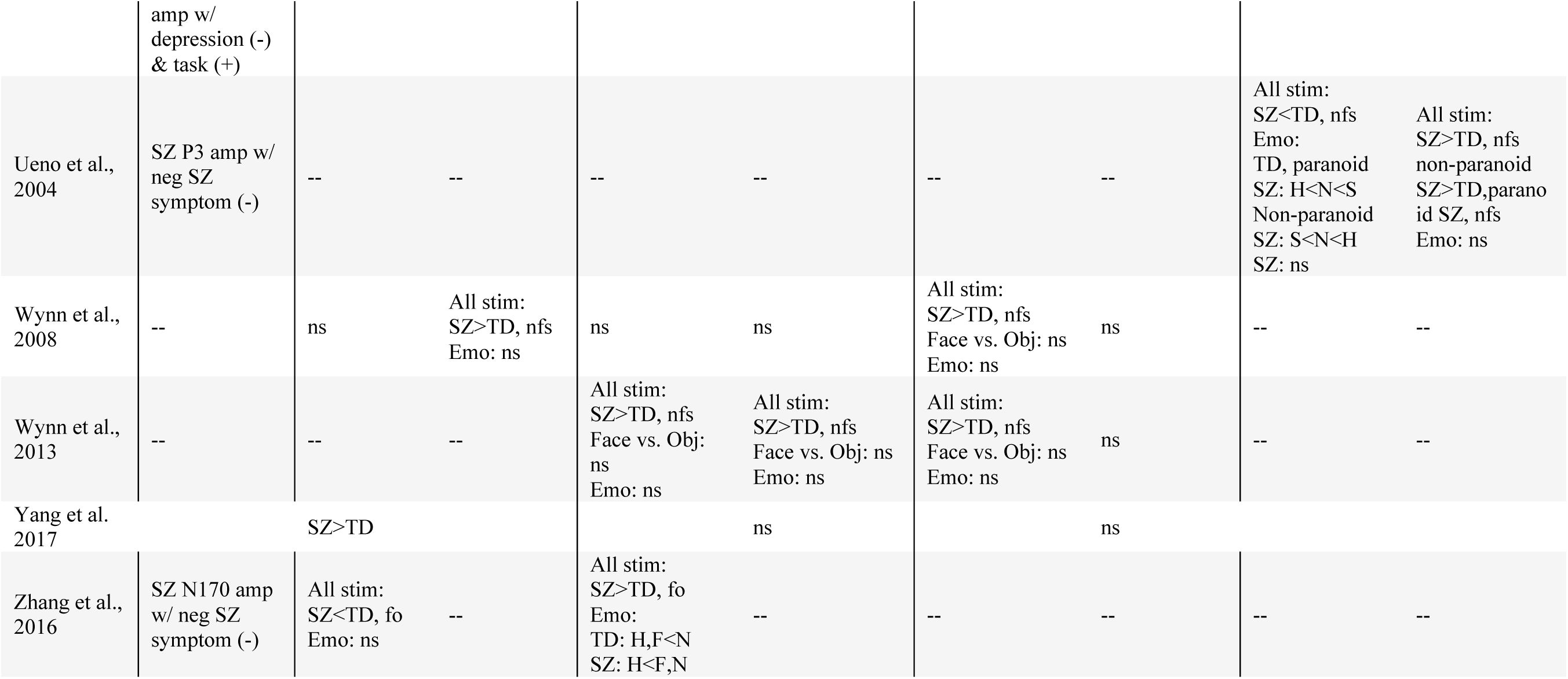
ERP results from all reviewed studies. Angry (A), Attention Deficit-Hyperactivity Disorder group (ADHD), amplitude (amp), Autism Spectrum Disorder group (ASD), disgusted (D), emotion (emo), fear (F), face-only effect (fo; study does not include non-face stimuli), happy (H), latency (lat), ashamed (M), neutral (N), negative (neg), not face-specific effect (nfs; effect across face and non-face stimuli), object (obj), positive (pos), surprised (R), sad (S), schizophrenia group (SZ), typically developing group (TD), positive correlation (+; larger magnitude or faster latency correlated with greater performance or fewer symptoms), negative correlation (-; larger magnitude or faster latency correlated with poorer performance or more symptoms. Note: for negative-going components, smaller amplitude indicates greater magnitude.

**Fig 1.**
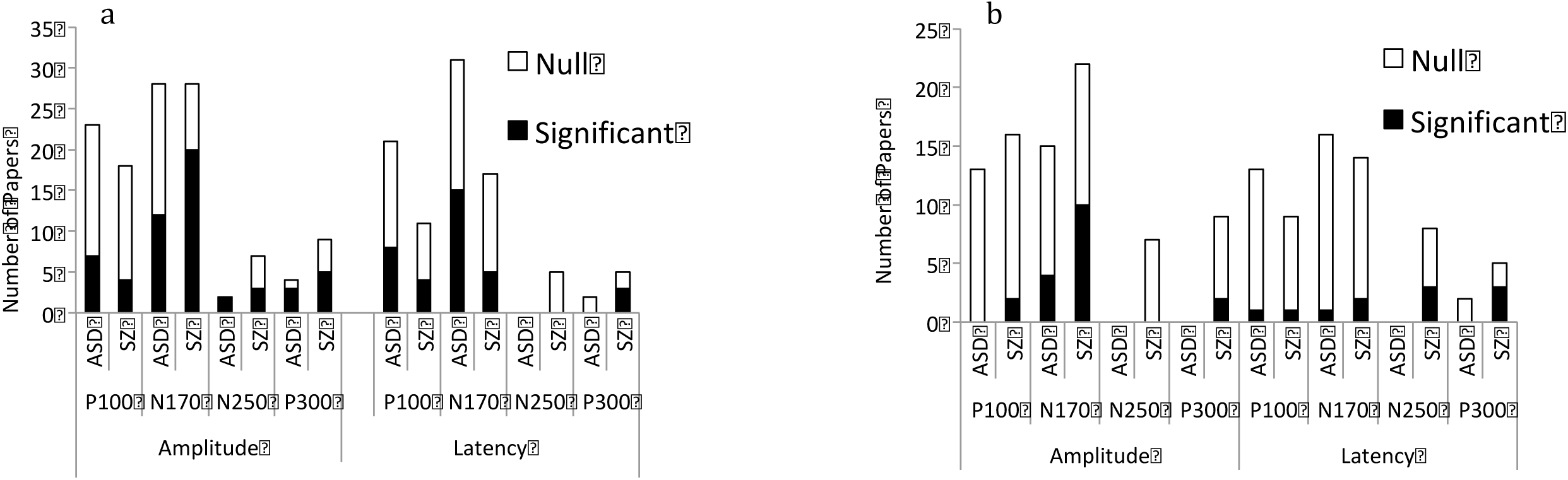
Number of studies reporting significant versus null findings across all studies, with all face processing effects summarized in Figure 1a and all emotion-related effects summarized in Figure 1b. Latency (lat), amplitude (amp), significant differences between groups (sig), comparable values between groups (null)

### P100 Amplitude

#### P100 Amplitude in ASD: Face

The majority of ERP literature reported no significant differences in P100 amplitude to faces between individuals with ASD and TD. Twenty-three studies of face and emotion processing in ASD reported on P100, 16 of which reported comparable P100 amplitudes in ASD and TD. In these 16 studies, no significant group differences in P100 amplitude to faces were identified across a range of task demands (attending to non-social target images to actively attending to facial stimuli) and in both child and adult samples (Akechi et al. 2010; Webb et al. 2010; Tye et al. 2013; Senju et al. 2005; Wagner et al. 2013; J. C. McPartland et al. 2011; Apicella et al. 2013; O’Connor et al. 2005 [child and adult]; 2007; T. K. Wong et al. 2008; Neuhaus et al. 2016; Shen et al. 2016; Yamasaki et al. 2017; Luyster et al. 2017[child and adult]).

Studies reporting group differences (7 of 23) in P100 amplitude to faces revealed inconsistent directionality of effects. One study of adults found increased P100 amplitude to faces in ASD versus TD (Cygan et al. 2014). One child study and one adult study reported inversion effects (greater P100 amplitude to inverted versus upright faces) in the TD group, but not in the ASD group (Hileman et al. 2011; Webb et al. 2012). Poorer face memory in Webb and colleagues’ study (2012) was correlated with greater P100 response to inverted versus upright faces in adults with ASD; however, there were no significant differences between groups in processing upright faces or objects. One study of children reported attenuated P100 amplitude to faces in ASD compared to TD, but lacked control stimuli (M. Batty et al. 2011), and a second study replicated this effect in young adults when using photographs of emotional faces, but reported a slightly larger P100 amplitude in ASD when using realistic drawings of emotional faces (Tseng et al. 2015). Tseng’s study reported no differential effect of each emotion on the P100 response within or across groups; thus, this is a face-related, rather than emotion-specific, result. An additional study of children reported reduced P100 amplitude in ASD for repeated versus novel faces and objects whereas no difference was found in TD (A. P. Key and Corbett 2014). A final study found reduced P100 lateralization in the ASD group as compared to controls (Luckhardt et al. 2017). Of these seven studies, three found effects across face and objects, and four did not include non-face stimuli. As such, no studies found a face-stimulus specific effect.

#### P100 Amplitude in ASD: Emotion

There were 13 studies of emotion processing in ASD that reported on P100 amplitude, distributed across childhood, adolescence, and adulthood. Across all studies, P100 was modulated similarly between diagnostic groups in response to emotional faces (Apicella et al. 2013; Akechi et al. 2010; M. Batty et al. 2011; Hileman et al. 2011; Luyster et al. 2017 [child and adult]; O’Connor et al. 2005 [child and adult]; 2007; Wagner et al. 2013; T. K. Wong et al. 2008; Luckhardt et al. 2017; Yamasaki et al. 2017).

#### P100 Amplitude in SZ: Face

Similar to findings in ASD, most studies of face processing (14 of 18) reported comparable P100 amplitudes to face stimuli between SZ and TD groups across a variety of methodologies and sample compositions (Bediou et al. 2007; Kim et al. 2015; Colleen A Brenner et al. 2015; Fukuta et al. 2014; Herrmann et al. 2004; Jetha et al. 2013; P. J. Johnston et al. 2005; Jung et al. 2012; S.-H. Lee et al. 2010; Obayashi et al. 2009; Thoma et al. 2013; Turetsky et al. 2007; Wynn et al. 2008; Liu et al. 2016). Among studies reporting P100 amplitude differences, three found attenuated P100 to faces in SZ groups (Caharel et al. 2007; Campanella et al. 2006; Zhang et al. 2016), though Campanella (2006) found that this effect was specific to a group displaying high SZ symptomatology (compared to TD and low-symptom SZ groups). One study found the opposite effect, observing larger P100 amplitude to faces in the SZ group (Yang et al. 2017). In all four of the studies that found significant differences, participants were actively attending to facial stimuli; twelve of the fourteen studies that found null results also included active attention to stimuli. Importantly, only 2 of 18 studies included control (i.e., non-face) stimuli, and neither of these studies reported differences between SZ and TD groups (Obayashi et al. 2009; Wynn et al. 2008).

#### P100 Amplitude in SZ: Emotion

Out of 16 emotion-processing studies reporting P100 amplitude in SZ, 14 studies reported no significant group differences in P100 amplitude as a function of the emotional content of presented faces (P. J. Johnston et al. 2005; Fukuta et al. 2014; Colleen A Brenner et al. 2015; Caharel et al. 2007; Kim et al. 2015; Campanella et al. 2006; Jung et al. 2012; S.-H. Lee et al. 2010; Obayashi et al. 2009; Thoma et al. 2013; Turetsky et al. 2007; Wynn et al. 2008; Zhang et al. 2016; Yang et al. 2017). One study reported amplitude modulation by emotion in the TD group but not in the SZ group (Bediou et al. 2007), and a second reported reduced P100 amplitude in SZ compared to controls for disgust, but not fear or happy faces (Yang et al 2017). Jetha and colleagues (2013) reported that moderate shyness in SZ was associated with less emotional reactivity as indicated by P100 amplitude. This finding suggests that differences in P100 amplitude may mark alterations in emotion processing associated with specific symptom dimensions. Taken together, these results suggest that, as in the ASD literature, P100 amplitude does not specifically index alterations in face or emotion processing.

### P100 Latency

#### P100 Latency in ASD: Face

Almost two-thirds of studies (13 of 21) did not find significant differences between ASD and TD groups regarding P100 latency to faces (J. C. McPartland et al. 2011; O’Connor et al. 2005 [child]; Apicella et al. 2013; Cygan et al. 2014; Hileman et al. 2011; O’Connor et al. 2007; Wagner et al. 2013; T. K. Wong et al. 2008; Senju et al. 2005; Webb et al. 2010; Webb et al. 2012; Luckhardt et al. 2017; Luyster et al. 2017 [child]). Eight studies reported group differences in response to facial stimuli. Two child and two adult studies reported increased latency to faces in ASD groups compared to the TD group (M. Batty et al. 2011; O’Connor et al. 2005 [adult]; Neuhaus et al. 2016; Luyster et al. 2017 [adult]). Of note, both Luyster (2017) and O’Connor et al.’s (2005) child sample did not show differences between groups despite using the same methods as in their adult samples, where P100 latency delays were identified.. A single study found shorter P100 latencies to faces in adults with ASD versus TD (Yamasaki et al. 2017). Two other child studies reported group differences in lateralization, wherein P100 latency was faster in the right versus left hemisphere in TD, but this effect was absent in ASD (Akechi et al. 2010; Tye et al. 2013). A final study showed shorter left hemisphere P100 latencies present only in the ASD group (Shen et al. 2016).

#### P100 Latency in ASD: Emotion

Twelve of thirteen studies of emotion processing in ASD did not report significant group differences between TD and ASD groups in P100 latency sensitivity to emotion (Akechi et al. 2010; Apicella et al. 2013; Hileman et al. 2011; Luckhardt et al. 2017; Luyster et al. 2017 [child and adult]; O’Connor et al. 2005 [child and adult]; 2007; Wagner et al. 2013; T. K. Wong et al. 2008; Yamasaki et al. 2017). However, Batty et al. (2011) reported that P100 was modulated by emotion in TD children, but not in ASD.

#### P100 Latency in SZ: Face

Seven of 11 studies reporting P100 latency to face stimuli reported no significant differences between SZ and TD groups (Herrmann et al. 2004; Campanella et al. 2006; Fukuta et al. 2014; Kim et al. 2015; P. J. Johnston et al. 2005; Thoma et al. 2013; Liu et al. 2016). The remaining studies reported longer P100 latency in SZ groups compared to TD groups (Caharel et al. 2007; Obayashi et al. 2009; S.-H. Lee et al. 2010; Wynn et al. 2008). None of these results are clearly face-specific, however; two studies did not include control stimuli (Caharel et al. 2007; S.-H. Lee et al. 2010), and two found P100 latency delays in SZ across both face and control stimuli (Obayashi et al. 2009; Wynn et al. 2008).

#### P100 Latency in SZ: Emotion

Out of nine studies, eight reported no difference between P100 latency in SZ and TD groups in response to emotional faces (Caharel et al. 2007; Campanella et al. 2006; Fukuta et al. 2014; P. J. Johnston et al. 2005; Kim et al. 2015; Obayashi et al. 2009; Thoma et al. 2013; Wynn et al. 2008). One study reported sex-specific diagnostic differences in emotion processing: in females, P100 in the left hemisphere was delayed in SZ compared to TD when attending to happy faces (S.-H. Lee et al. 2010). Moreover, longer latency to happy faces correlated with more negative symptomatology in SZ females. In SZ males, there was no overall delay in P100 compared to the TD group, and longer latency correlated with better performance on a behavioral detection task for happy versus fear faces.

### N170 Amplitude

#### N170 Amplitude in ASD: Face

Since the initial report of N170 as a marker of atypical social perception in ASD (J. McPartland et al. 2004), 28 studies have examined N170 amplitude to faces in individuals with ASD, with 12 reporting differences in face processing by diagnosis. Of those 12, six studies (two in children and four in adults) reported attenuated amplitude in ASD compared to TD (Cygan et al. 2014; O. Churches et al. 2012b; O’Connor et al. 2005 [adult]; Tye et al. 2014; Webb et al. 2012; Groom et al. 2017). Six other studies reported differences in N170 response to faces between ASD and TD groups, but these differences were not always evident in direct statistical comparisons between groups (e.g., absence of lateralization in one group without demonstration of a group by lateralization interaction). One such study of children reported a typical face inversion effect in TD but the opposite effect in ASD (J. C. McPartland et al. 2011). Three studies reported right hemisphere lateralization in TD but lack of lateralization in ASD (J. C. McPartland et al. 2011; Tye et al. 2013; Webb et al. 2006). Three studies reported findings of N170 amplitude differentiation between: face stimulus types in TD but not ASD (Akechi et al. 2010), active versus passive tasks in TD but not ASD (Owen Churches et al. 2010), and repeated versus novel faces and objects in ASD but not TD (A. P. Key and Corbett 2014).

Most studies reporting correlative analyses demonstrated associations between N170 amplitude and autistic symptomatology or social behavior. More negative N170 amplitude to inverted faces correlated with better face recognition in children with ASD (J. C. McPartland et al. 2011) and better social skills in adults with ASD (Webb et al. 2012). Stronger overall N170 amplitude to faces was associated with more typical social behavior in TD children (though not in the ASD group; (Hileman et al. 2011), and increased right lateralization was associated with better social communication across both TD and ASD children (Tye et al. 2013).

Sixteen of the 28 studies of N170 amplitude to faces in ASD (1 of children, 5 distributed across childhood into adolescence, 2 of adolescents, 2 spanning adolescence into early adulthood, and 6 of adults) did not find differences between diagnostic groups (Khorrami et al. 2013; O. Churches et al. 2012a; Grice et al. 2005; Wagner et al. 2013; J. McPartland et al. 2004; Apicella et al. 2013; T. K. Wong et al. 2008; M. Batty et al. 2011; Senju et al. 2005; O’Connor et al. 2007; Magnée et al. 2011; O’Connor et al. 2005 [child]; Hileman et al. 2011; Webb et al. 2010; Faja et al. 2016; Luckhardt et al. 2017). Approximately half of these studies did, however, find differences in N170 latency, reported below. Of note, most of these studies did not instruct participants to attend to the faces specifically but included non-face targets to monitor attention. Only four of 16 studies with null results required participants to attend to faces. In contrast, all but one study (Webb et al. 2010) that reported differences between ASD and TD groups required participants to actively attend to stimuli. Given evidence that N170 amplitude is enhanced by attention (Owen Churches et al. 2010; Mohamed et al. 2009), this finding suggests that differentiation of N170 amplitude emerges when the task demands are most face-specific.

#### N170 Amplitude in ASD: Emotion

Eleven of 15 studies reported no significant difference in N170 amplitude between ASD and TD groups during emotional face processing (Apicella et al. 2013; M. Batty et al. 2011; Hileman et al. 2011; Magnée et al. 2011; O’Connor et al. 2005 [child and adult]; 2007; T. K. Wong et al. 2008; Luckhardt et al. 2017; Luyster et al. 2017[child and adult]). The remaining four studies reported group differences. In two of these studies, N170 amplitude was modulated by emotion in TD groups but not in ASD groups (Akechi et al. 2010; Wagner et al. 2013). One study found the opposite effect, showing N170 amplitude modulation by emotion only in the ASD group (Faja et al. 2016). This last finding is consistent with one other finding in which ASD groups with and without comorbid ADHD showed increased N170 amplitude to neutral relative to fearful faces, whereas amplitude in the TD group was not modulated by emotional valence (Tye et al. 2014). More negative N170 amplitude overall, as well as more negative amplitude to fearful compared to neutral faces, correlated with better social communication skills across both TD and ASD children (Tye et al. 2014). Contrary to patterns detected among face processing findings, only one of these four studies yielding group differences required participants to attend to the emotional valence of the stimuli; the other two required participants to attend to non-social target stimuli. Likewise, four studies with null results tasked participants with identifying the emotional valence of stimuli (O’Connor et al. 2005 [child and adult]; 2007; T. K. Wong et al. 2008). Thus, the degree to which explicit demands for attention to emotion may contribute to N170 amplitude alteration in ASD for emotion processing is less clear than for face stimuli.

#### N170 Amplitude in SZ: Face

Twenty out of 28 studies investigating N170 amplitude to faces in adults with SZ reported significant differences in face processing between diagnostic groups. In four of these studies, amplitude to faces was smaller in SZ relative to TD, whereas amplitude to objects was not significantly different between groups (Herrmann et al. 2004; Obayashi et al. 2009; Onitsuka et al. 2006; Tsunoda et al. 2012). One study observed N170 attenuation in SZ across inverted face and non-face images (Zheng et al. 2016). Another study showed right lateralization of N170 to low-spatial frequency compared to high spatial frequency of fearful faces in SZ but no differentiation in TD groups (Kim et al. 2015). Fourteen additional studies report attenuated N170 amplitude to faces in SZ relative to TD; however, across all these studies, findings were not necessarily face-specific, as several lacked non-face control stimuli (Bediou et al. 2007; Caharel et al. 2007; Campanella et al. 2006; Jetha et al. 2013; Jung et al. 2012; Kirihara et al. 2012; S.-H. Lee et al. 2010; Onitsuka et al. 2009; Turetsky et al. 2007; Zhang et al. 2016) and the remainder showed main effects of group where N170 amplitude attenuation spanned both face and non-face stimuli (Ibanez et al. 2012; Lynn and Salisbury 2008; Wynn et al. 2013; Liu et al. 2016).

A handful of studies reported correlations between greater N170 amplitude to faces and higher global functioning (Obayashi et al. 2009), less positive SZ symptomatology (Campanella et al. 2006), decreased shyness (Jetha et al. 2013), higher social competency (Tsunoda et al. 2012), and increased extraversion (Kirihara et al. 2012). Together, these studies provide strong evidence for an association between more robust N170 response to faces and better social and psychiatric functioning across both TD and SZ populations. Eight studies reported no significant differences in N170 amplitude to faces between diagnostic groups (Akbarfahimi et al. 2013; Colleen A Brenner et al. 2015; Komlosi et al. 2013; S. Lee et al. 2007; Ramos-Loyo et al. 2009; Tso et al. 2015; Thoma et al. 2013; Wynn et al. 2008). However, one found emotion-specific results that are detailed in the following section (Thoma et al. 2013), and the second found decreased N170 differentiation to facial angle as compared to TD controls (Tso et al. 2015).These eight studies have comparable methodologies and group demographics to those studies that do detect group differences.

#### N170 Amplitude in SZ: Emotion

A number of studies (10 of 22 suggest altered N170 amplitude during emotion discrimination in SZ compared to TD (Tso et al. 2015; Thoma et al. 2013; Caharel et al. 2007; Campanella et al. 2006; Lynn and Salisbury 2008; Jung et al. 2012; Turetsky et al. 2007; Zhang et al. 2016; Kirihara et al. 2012; Ibanez et al. 2012). Four of these studies reported that TD groups showed modulation of N170 amplitude based on emotional valence, whereas SZ groups did not (Campanella et al. 2006; Ibanez et al. 2012; Kirihara et al. 2012; Lynn and Salisbury 2008). Three studies reported attenuated N170 amplitude in SZ compared to TD to specific emotions, such as disgust, sadness, and happiness (Caharel et al. 2007; Jung et al. 2012; Turetsky et al. 2007). One found that N170 amplitude was differentially modulated by emotion in SZ relative to TD: whereas in TD it was larger for happy and fearful faces than for neutral faces, in SZ it was larger for happy faces than for neutral or fearful (Zhang et al., 2016). Additionally, reduced left hemisphere N170 amplitude to fearful faces in particular was correlated with severity of blunted affect in patients. Two additional studies found group differences as a function of more nuanced influences on emotion perception, including larger left lateralized N170 amplitude in fearful vs. neutral averted-gaze faces in SZ (Tso et al. 2015), and enhanced N170 amplitude in TD, but not SZ, when the emotion displayed in a stimulus differed from the stimulus emotion proceeding it (Thoma et al. 2013).

Twelve studies investigating N170 amplitude reported no significant differences between SZ and TD in this ERP component during emotion processing (Jetha et al. 2013; Bediou et al. 2007; Colleen A Brenner et al. 2015; S.-H. Lee et al. 2010; Obayashi et al. 2009; Kim et al. 2015; Wynn et al. 2013; Akbarfahimi et al. 2013; Komlosi et al. 2013; S. Lee et al. 2007; Ramos-Loyo et al. 2009; Wynn et al. 2008). These studies have comparable population demographics and methodologies to the studies above where differences were detected.

### N170 Latency

There were 31 studies of ASD that reported N170 latency. Eighteen of these studies were performed in children, and 13 were in adults. Findings below are separated by age.

#### N170 Latency in ASD: Face

Of the 18 studies investigating N170 latency during face processing in children with ASD, eight reported atypical temporal processing in the ASD group. One study reported face-specific delays in the ASD group compared to the TD group (J. C. McPartland et al. 2011), another reported delayed latency to faces compared to objects in the ASD group, contrasted with faster latency to faces than objects in the TD group (Webb et al. 2006), and four studies found slower N170 latency in ASD compared to TD groups across both face and non-face stimuli (Khorrami et al. 2013; M. Batty et al. 2011; Hileman et al. 2011; Luyster et al. 2017 [child]). A study by Gunji et al (2009) found these results when comparing a child ASD group to adult controls but not to child controls. One study observed shorter N170 latencies in the left hemisphere only in the ASD group across both social and nonsocial stimuli, (Shen et al. 2016); this finding is in the opposite direction than others, indicating faster N170 latency in ASD. Ten studies in children did not find significant between-group differences in N170 latency to faces (Akechi et al. 2010; Tye et al. 2014; Apicella et al. 2013; O’Connor et al. 2005 [child]; Senju et al. 2005; Grice et al. 2005; Tye et al. 2013; Wagner et al. 2013; T. K. Wong et al. 2008; Luckhardt et al. 2017). There were no observable differences in methodology or group characteristics to account for variations in study findings.

In adults, seven of 13 studies indicated atypical N170 latency in ASD groups. Across two studies, faces elicited a slower N170 latency in ASD compared to TD, while between-group differences in N170 latency to object images were not significant (J. McPartland et al. 2004; O’Connor et al. 2007); an additional three studies also found N170 delays to faces, but there were no non-social control stimuli in these experiments (O’Connor et al. 2005 [adult]; Yamasaki et al. 2017; Luyster et al. 2017 [adult]). Two studies suggest a relation between N170 latency and clinical symptoms. McPartland et al (2004) found that, in ASD but not controls, slower N170 latency to both upright and inverted faces associated with better face memory, suggesting that longer processing times may confer improved behavioral performance. Though Webb et al (2012) did not find deficits in in N170 latency to upright faces relative to either houses or inverted faces or replicate McPartland et al’s (2004) association between N170 latency with face memory, faster N170 to upright *versus* inverted faces was related to both better social skills and better face memory. Two studies reported that topographic differences in N170 latency between ASD and TD. N170 latency to faces was faster medially than laterally in TD (Webb et al. 2012), but trending towards faster laterally than medially in ASD (Webb et al. 2010). Finally, six studies of adults with ASD found no significant between-group differences or differences in lateralization in N170 latency to face stimuli (O. Churches et al. 2012a; O. Churches et al. 2012b; Magnée et al. 2011; Owen Churches et al. 2010; Cygan et al. 2014; Tseng et al. 2015). A recent meta-analysis of face processing studies in ASD indicated that, on average, N170 latency was delayed in ASD relative to TD (Kang et al. 2018).

#### N170 Latency in ASD: Emotion

Fifteen of sixteen studies of children and adults converge in reporting no emotion-dependent differences in N170 latency between individuals with ASD and TD (Akechi et al. 2010; Magnée et al. 2011; Apicella et al. 2013; M. Batty et al. 2011; Hileman et al. 2011; O’Connor et al. 2005 [child and adult]; 2007; Wagner et al. 2013; T. K. Wong et al. 2008; Luckhardt et al. 2017; Tseng et al. 2015; Yamasaki et al. 2017; Luyster et al. 2017 [child and adult]). Seven of these studies were of children only, two studies included both child and adult groups, and four studies only included adults. Just one study reported significant differences in emotion perception: when taking ADHD diagnosis into account, Tye and colleagues (2014) found a group by emotion interaction in which children with ASD showed a longer N170 latency to angry faces than to neutral faces and a shorter latency to happy faces than to fearful faces, whereas children with ASD and comorbid ADHD showed the opposite pattern and TD children did not show any differences in N170 latency across emotions. This finding suggests that a cross-diagnostic approach to identifying neural indices of emotion processing may be more informative than studies restricted to one clinical group.

#### N170 Latency in SZ: Face

Twelve of 17 studies reported no difference in N170 latency between diagnostic groups in response to face stimuli (Campanella et al. 2006; Tso et al. 2015; Herrmann et al. 2004; Tsunoda et al. 2012; S. Lee et al. 2007; P. J. Johnston et al. 2005; Kim et al. 2015; Lynn and Salisbury 2008; Obayashi et al. 2009; Thoma et al. 2013; Wynn et al. 2008; Yang et al. 2017). The remaining five studies found adults with SZ showed delayed N170 latency to faces compared to TD controls. One of these studies found these results for both upright and inverted faces (Zheng et al 2016) and showed that N170 latency correlated with both negative and general psychiatric symptoms in ASZ patients. However, because the other four studies either had no control stimuli (Caharel et al. 2007; Wynn et al. 2013; S.-H. Lee et al. 2010) or saw main effects across both face and non-face stimuli (Akbarfahimi et al. 2013; S.-H. Lee et al. 2010; Caharel et al. 2007), N170 delays in SZ are not necessarily face-specific.

#### N170 Latency in SZ: Emotion

Twelve of 14 studies of emotion processing reported no significant differences between SZ and TD in N170 latency modulation to emotional faces (Campanella et al. 2006; Caharel et al. 2007; S. Lee et al. 2007; P. J. Johnston et al. 2005; Kim et al. 2015; Tso et al. 2015; Lynn and Salisbury 2008; Obayashi et al. 2009; Thoma et al. 2013; Wynn et al. 2008; Wynn et al. 2013; Yang et al. 2017). Two studies reported a differential effect of emotional faces on N170 latency in SZ versus TD. In a passive viewing study, adults with SZ showed a delayed N170 to fearful faces relative to other emotional faces, whereas controls displayed the shortest latencies to fearful faces (Akbarfahimi et al. 2013). Investigating gender differences, Lee et al. (2010) reported emotion-specific differences between females, but not males, with SZ and TD: in the right hemisphere, happy and fearful faces elicited a delayed N170 in females with SZ compared to those with TD. Slower N170 latency to happy faces in females with SZ was also correlated with increased expression of negative SZ symptomatology. An earlier study from the same group reported faster N170 latency to happy and neutral faces was associated with increased positive and negative SZ symptomatology, but found no significant differences between groups (S. Lee et al. 2007); thus, examining gender-specific effects may clarify the nature of processing deficits and relations with clinical symptoms.

### N250 Amplitude

#### N250 Amplitude in ASD: Face

Two studies in adults with ASD reported on N250 amplitude, and both found significant, yet conflicting, group differences. When viewing a series of neutral faces and instructed to attend to a target face, relative to TD adults, adults with ASD showed reduced N250 amplitude to targets (O. Churches et al. 2012a). Groups did not differ in their N250 response to non-target faces. In contrast, Webb et al. (2010) reported enhanced N250 amplitude to both familiar and unfamiliar face stimuli in the ASD group relative to TD adults. In the same study, N250 amplitude difference between familiar versus unfamiliar faces was right lateralized in ASD, while it was uniform across hemispheres in the TD group. Differing methodologies across these two studies may have elicited different main effects. The first used unfamiliar target faces in an active viewing task demanding attention and facial memory; the second used familiar faces in a passive viewing task. Of note, (Webb et al. 2010) also reported that increased right lateralization of N250 to familiar versus unfamiliar faces in the ASD group correlated with lower ADOS scores in communication and social domains.

#### N250 Amplitude in ASD: Emotion

There were no studies of emotion processing in ASD that reported N250 amplitude.

#### N250 Amplitude in SZ: Face

Three of seven studies of SZ measuring N250 amplitude to faces reported group differences. Two of these studies reported attenuated N250 in SZ groups relative to TD groups to both face and non-face stimuli (Wynn et al. 2013; Wynn et al. 2008). These studies used the same methodology and the authors noted that while their TD groups were different across the two studies, their SZ groups were highly overlapping. A third investigated sex differences and reported larger N250 amplitudes in males versus females with SZ when attending to faces; this sex difference was absent in TD (Jung et al. 2012). Overall, however, no between group (SZ versus TD) differences were significant. The remaining four studies did not find significant differences in N250 amplitude between groups (S.-H. Lee et al. 2010; Kim et al. 2015; Colleen A Brenner et al. 2015; Turetsky et al. 2007).

#### N250 Amplitude in SZ: Emotion

Of the seven studies reported above, all used emotional stimuli, but none found group differences in N250 amplitude as a function of the emotional valence of faces.

### N250 Latency

#### N250 Latency in ASD

There were no studies of face or emotion processing in individuals with ASD that reported N250 latency.

#### N250 Latency in SZ

There were five studies of face and emotion processing in SZ that reported N250 latency; however, none found significant group differences, either to faces overall or to emotional faces specifically (S.-H. Lee et al. 2010; Kim et al. 2015; Wynn et al. 2013; Wynn et al. 2008; Yang et al. 2017).

### P300 Amplitude

#### P300 Amplitude in ASD: Face

Three of four studies of ASD reporting on P300 amplitude detected differences between diagnostic groups, albeit in conflicting directions. In two studies, familiar faces elicited greater P300 than unfamiliar faces in TD children, whereas this effect was absent in children with ASD (Gunji et al. 2009; Gunji et al. 2013). In contrast, (A. P. Key and Corbett 2014) found enhanced P300 amplitude to repeated faces in children with ASD relative to the TD group. P300 amplitude was also greater to repeated faces than to neutral stimuli in ASD but not TD. However, in this study, the same pattern was seen for repeated objects, suggesting the enhanced P300 effect during stimulus repetition was not necessarily face-specific. When measuring neural response to averted and direct eye gaze, there were no significant differences in P300 amplitude between children with ASD and TD to either stimulus type (Senju et al. 2005).

#### P300 Amplitude in ASD: Emotion

There were no studies of emotion processing in ASD that reported P300 amplitude.

#### P300 Amplitude in SZ: Face

Five of nine studies of SZ reporting on P300 amplitude detected diagnostic differences during reception of facial stimuli. In three of these studies, P300 amplitude was attenuated in SZ relative to TD, although these results were not face-specific (Ramos-Loyo et al. 2009; Turetsky et al. 2007; Ueno et al. 2004). However, greater P300 amplitude was correlated with lower negative SZ symptomatology (Ueno et al. 2004) and better emotion identification (Turetsky et al. 2007), suggesting its relevance for social symptomatology specifically. In a study of sex differences, while P300 amplitude was equivalent in TD males and females, females with SZ showed a larger P300 amplitude to faces than males with SZ; overall, there were no between group differences with regards to P300 amplitude (S.-H. Lee et al. 2010). Finally, a study investigating effects of anti-psychotic medication reported that, relative to TD controls, the SZ group showed attenuated P300 amplitude to faces, but only after exposure to 12 months of medication (Mori et al. 2012). Four studies reported no group differences in P300 amplitude to faces (Bediou et al. 2007; Kim et al. 2015; P. J. Johnston et al. 2005; Jung et al. 2012). There were no clear patterns among results to indicate methodological or sample differences accounting for this variability.

#### P300 Amplitude in SZ: Emotion

Two of nine studies of emotion processing reported significant group differences in P300 amplitude to emotional faces in SZ. When observing images of infant faces and objects, TD adults showed the largest P300 amplitude to a crying infant face and the smallest amplitude to a happy face (Ueno et al. 2004), suggesting that an infant’s distress is most salient to TD adults. In the SZ group as a whole, P300 amplitude was not modulated by the emotional valence of the infant face; however, when the SZ group was divided by subtype, adults with paranoid SZ showed the same pattern as the TD group, while those with non-paranoid SZ showed an inverted effect, with larger P300 amplitude to happy than to crying faces. In a second study, fearful adult faces elicited greater P300 amplitude than neutral faces in TD adults tasked with attending to emotional valence. There was no modulation of the P300 response by emotion in the SZ group (Bediou et al. 2007). The remaining seven studies found no significant group differences in emotion processing as measured by P300 amplitude (P. J. Johnston et al. 2005; Kim et al. 2015; Mori et al. 2012; Ramos-Loyo et al. 2009; S.-H. Lee et al. 2010; Turetsky et al. 2007; Jung et al. 2012).

### P300 Latency

#### P300 Latency in ASD

Two face processing studies in ASD reported on P300 latency; neither found group differences (Gunji et al. 2013; Senju et al. 2005). No emotion processing studies in ASD reported on P300 latency.

#### P300 Latency in SZ

Three of five studies of face and emotion processing that reported on P300 latency in SZ found group differences. Two studies found delayed P300 in SZ compared to TD in response to all stimuli, regardless of whether stimuli were faces or objects, or of the emotional valence of face stimuli (S.-H. Lee et al. 2010; Ueno et al. 2004). Latency differences were, however, modulated by paranoia. Specifically, adults with non-paranoid SZ showed longer P300 latency to faces than TD adults, whereas individuals with paranoid SZ did not differ from the TD group (Ueno et al. 2004). This pattern parallels the P300 amplitude finding from the same study. In a third study, adults with SZ who had been medicated for 12 months showed delayed P300 latency to happy—but not neutral or sad—faces relative to TD adults (Mori et al. 2012). In the same study, a slowing effect of medication on P300 latency in SZ was apparent: adults with SZ showed slower P300 latency after 12 months of medication compared to their responses while medication-naïve and medicated for a shorter, 3-month interval. Two studies did not report group differences in P300 latency (P. J. Johnston et al. 2005; Kim et al. 2015).

## DISCUSSION

Here we have reviewed the results of studies reporting on P100, N170, N250, and P300 ERP components in response to neutral and emotional face stimuli in individuals with autism spectrum disorders and schizophrenia. Our review sheds light on patterns of replicability in the literature and highlights multiple areas marked by inconsistent findings and in need of further research.

For the P100, which reflects early stages of visual processing, there is little evidence of amplitude or latency alterations in ASD across face or emotion processing. Scattered findings suggest that P100 amplitude may index neural processes relevant to ASD, but primary P100 deficits are unlikely. As with face processing, P100 latency does not seem to be a reliable indicator of emotion processing differences between ASD and TD groups. While P100 latency may index variability in speed of low-level visual processing across groups, it does not capture behavioral differences in face or emotion processing between ASD and TD groups. In schizophrenia, there is similarly little evidence for P100 amplitude or latency alterations being effective indices of face or emotion processing deficits.

The N170, a well-studied index of face processing, was the component most consistently demonstrating atypical neural response across the two neurodevelopmental disorders. In ASD studies, approximately 40% of all findings reported diminished amplitude and delayed latency to faces in ASD compared to TD; however, several of these studies did not include non-face control stimuli or found effects that were not specific to faces. Two-thirds of the studies that found significant differences required active attention to stimuli, whereas three-quarters of the studies that found no significant differences were passive viewing tasks. This distinction suggests that N170 deficits in ASD are particularly prominent when attentional demands are involved. There was little evidence of N170 amplitude or latency reliably indexing deficits in in emotion processing in ASD. In studies of SZ, evidence for N170 latency delays to faces and differential N170 latency by emotion was mixed. However, there was strong evidence of diminished N170 amplitude to both face and non-face stimuli, indicating differences by diagnosis in early visual – but not necessarily face-specific – processing. There was also strong evidence of diminished N170 amplitude in SZ from studies of emotion processing. In general, N170 amplitude was differentially responsive to neutral versus emotionally-valenced faces in TD groups, whereas SZ groups did not tend to show this specialization. This pattern may suggest that processing the emotional content of faces, rather than just faces themselves, may be most specifically atypical in SZ. In contrast, early structural processing of faces in general, more so than differentiation of particular emotions, may be more consistently impacted in ASD.

Across the N250 and P300 components, there are too few studies in ASD to draw conclusions despite promising group differences. Further research comparing N250 and P300 amplitude in ASD and TD, along with re-analysis of previously published studies that did not include these components in their original analysis, may help elucidate the neural underpinnings of atypical face and emotion perception. Many studies that measured N250 and P300 amplitude found group differences in face perception, indicating that both components may be understudied but promising leads for indexing social cognition deficits.

In SZ, there is a slightly larger literature, but evidence for diminished N250 and P300 amplitude to face stimuli is mixed. N250 amplitude was attenuated to face, but not emotion, stimuli in approximately 40% of SZ studies, but N250 latency appears to be unaffected. P300 amplitude to face stimuli appear to be more reliably affected, and P300 amplitude may both mark symptom levels and be affected by medication treatments. Though findings from a small sample of studies suggest P300 latency may reliably index cognitive processing deficits for both face and emotion stimuli in SZ, the relative paucity of literature measuring P300 latency makes it difficult to identify this component as a sensitive neural marker of social cognition differences.

To expand both ASD and SZ literatures, given that ongoing, continuous EEG is recorded before segmentation for ERP analyses, it is likely that pre-existing data from studies reporting on earlier components (e.g., P100 and/or N170) could provide an opportunity to analyze N250 and P300 components to test for group differences during face and emotion processing. Such re-analysis of existing data may provide a quick and efficient mechanism to address outstanding questions of whether later, attentional neural responses are impacted in the context of reception of facial communication, particularly in ASD where existing data is more limited.

### Accounting for Variability Across Studies

Variation in methodologies – such as stimulus design and subject factors – may account for some of the inconsistencies among studies. Differences in low-level visual characteristics, such as luminance, contrast, and color, can produce significantly different waveforms (Bruno Rossion and Caharel 2011; Rousselet and Pernet 2011), yet most studies did not provide specific details on the low-level visual aspects of their stimulus set. Likewise, there were many different types of stimuli across studies. For example, while all studies included face stimuli, the nature of these faces varied (i.e., upright vs. inverted, familiar vs. unfamiliar, neutral vs. emotional, child vs. adult). Moreover, faces were sometimes the sole stimuli within a study paradigm, whereas other studies included non-face conditions. Study design, such as the use of blocking by stimulus type (e.g., face vs. non-face; neutral vs. emotional faces) vs. randomly interspersing all stimuli, can affect ERPs due to repetition and habituation effects (Shlomo Bentin and Peled 1990; Ravden and Polich 1999), which few studies examined. Importantly, many studies claiming group differences in face processing did not include non-face control stimuli, preventing conclusions regarding the specificity of reported findings to faces.

Participant tasks varied widely across studies, from passive viewing, to providing button-press responses to occasional attentional control trials not included in later analyses, to providing task-relevant verbal responses to each trial. Variability in task design yields inconsistencies in the nature and burden of the cognitive load placed on the participant and may manipulate how intensely and/or consistently the participant is attending to the stimuli. Unfortunately, there were very few instances in which clear patterns of replicable vs. less consistent findings were revealed as a function of particular aspects of stimulus type, task design, or demands for participant attention and task engagement. Finally, no studies in this review monitored participant gaze with eye-tracking; thus, while participants were ostensibly looking toward the screen (based on experimenter observations or attention monitoring tasks), gaze was not directly measured.

Subject factors also may have contributed to variability across studies. For example, not all studies matched clinical and control groups by IQ, making it difficult to ascertain which observed differences were due to diagnosis versus may have been driven by differences in cognitive functioning. Moreover, most studies did not account for sex when matching controls, and this variable may drive certain effects or correlations in ASD and SZ groups, as highlighted by several studies (Jung et al. 2012; S.-H. Lee et al. 2010). ERP components indexing face perception change in amplitude and latency over the lifespan (Roxane J Itier and Taylor 2004b; M. Batty et al. 2011), yet, when comparing studies of ASD and SZ, we are limited by the fact that most studies of ASD are in children, while all studies of SZ are in adults. Participants in clinical groups, especially SZ, were often medicated with psychoactive agents. In their longitudinal investigation of the effects of antipsychotic use on ERP waveforms, Mori and colleagues (Mori et al. 2012) reported changes in patients’ P300 amplitude and latency between drug-naïve and medicated time-points, and another study reported a positive correlation between medication dosage and P300 amplitude to emotional compared to neutral faces (Komlosi et al. 2013). Many studies did not report on the medication status of patients, and still others reported medication status but did not discuss testing for correlations between ERP waveforms and medication dose or controlling for medication status. Thus, effects of medication differences among participants and between clinical and control groups could also contribute to variability in findings across studies.

Variability in clinical profiles within participant samples could also have affected results. With respect to subject characterization, many studies did not report confirming ASD or SZ diagnoses in the context of the study with an ADOS (for ASD; Lord et al. 2012) or SCID (SCID, for SZ; First 1995). Lack of rigorous characterization of participants may mean that some clinical groups were not composed entirely of participants with gold-standard ASD or SZ diagnoses and/or may have included individuals falling short of diagnostic threshold. Many studies did not report level or severity or particular profiles of symptomatology within patient samples, despite the fact that ASD and SZ samples tend to be phenotypically heterogeneous. With disorders that are behaviorally-defined (and which likely lump individuals with multiple different etiological origins based on their symptoms; Happé et al. 2006; Insel 2010), re-grouping participants by subtypes or symptom profile can be a helpful way of finding patterns. Conducting correlations and regressions accounting for symptom levels on well-validated measures like the ADOS (for ASD; Lord et al. 2012) and Positive and Negative Syndrome Scale (PANSS, for SZ; Kay et al. 1987), as well as broader measures of symptoms that cut across diagnoses, may help further clarify the association between clinical profiles and neural processing abnormalities. For example, when dividing their SZ group into paranoid and non-paranoid subgroups, Ueno et al. (2004) found that the paranoid subgroup showed similar P300 amplitude and latency to the TD group, whereas the non-paranoid subgroup differed in P300 amplitude to emotional faces an in latency to all stimuli. When looking at the SZ group as a whole, however, these differences were obscured.

Related to the need to carefully characterize and account for variation in symptom profiles among participants is the need to recruit large study samples in order to capture heterogeneity and enable statistical analyses that are adequately powered to detect effects using covariates and regressions. Among the studies highlighted in this review, 50% of studies in ASD (18 of 36) and approximately 25% of those in schizophrenia (8 of 33) had fewer than 15 participants per group. Moreover, whereas nearly 25% of SZ studies (8 of 33) included 30 or more participants per group, only 5% of ASD studies (2 of 36) had sample sizes this large. Small samples are particularly vulnerable to spurious or non-generalizable findings (Button et al. 2013). Thus, it may be the case that some of the inconsistencies across studies relate to the relatively small sample sizes, particularly in the ASD literature. Moving forward, recruiting large samples to enable examination of more homogenous subsets of participants and to allow for evaluation of the association between ERP components and clinical traits measured in a continuous fashion might help to explain differences among studies where samples can be quite variable.

### Considerations for future research

This review of electrophysiological studies of face perception in ASD and SZ indicates that the N170, and possibly N250 and P300, hold promise as biomarkers indexing the integrity of social perceptual systems across neurodevelopmental disorders and for providing clues about underlying neural dysfunction within and across disorders. Several caveats for future research emerge from these findings.

First, this review makes clear that alterations in the highlighted ERP indices are not diagnostically specific. Similar patterns of atypical processing at identical components were observed in both ASD and SZ. This suggests that, consistent with the RDoC framework, it may be most useful to evaluate the underlying neural bases of neurodevelopmental disorders in terms of specific, dimensionally-measured processes related to function and dysfunction transdiagnostically, rather than designing studies around a particular diagnostic category.

Although the reviewed evidence does not make a reliable case that any one particular ERP component may be a consistent diagnostic biomarker, correlations with symptomatology and key associated features (e.g., emotion recognition) indicate promise with respect to other areas of biomarker development. These include classifying individuals into subgroups relevant to treatment selection (stratification biomarkers), demonstrating the influence of an intervention on a targeted neural system (target engagement biomarkers), or offering short-term indication of intervention effects on symptoms or underlying neural systems (early efficacy biomarkers; James C McPartland 2016). Our review focused specifically on ASD and schizophrenia; however, social cognition and these ERP components are germane to multiple other psychiatric diagnoses (Feuerriegel et al. 2015). Increased focus on transdiagnostic samples can provide greater clarity regarding the information these ERP indices provide about the neural processes underlying neurodevelopmental and psychiatric disorders.

As the variability in findings across existing studies makes clear, it is necessary to carefully consider individual differences in participant characteristics, such as age, both within and across diagnostic groups. Differences in findings reported within ASD between child and adult samples and cross-diagnostically between studies of ASD and SZ may reflect differences in age-related characteristics of particular samples. For example, some alterations in neural responses may be more prominent in adults, reflecting later onset difficulties or premature developmental plateaus, whereas some may be most notable in young children and resolve over time. Future research on markers of social perception must either constrain age ranges to reduce variability, conduct studies with sufficient statistical power to co-analyze age effects, or examine participant samples longitudinally to study the divergence of particular neural responses as they relate to emergence or amelioration of symptoms over time. Though age effects are less germane to schizophrenia, a disorder most commonly occurring in adulthood, whether alterations in ERP response to face stimuli are present in prodromal phases is an important question. Moreover, as other factors such as cognitive ability are also demonstrated to influence ERP indices, these characteristics must be considered and controlled, either statistically or through reduction of sample heterogeneity. Finally, prioritization of exploring hypotheses in large participant samples, either within individual studies or by harnessing “big data” sources, such as the National Institute of Mental Health’s RDoC or NDAR data repositories (data-archive.nimh.nih.gov), will maximize the probability of detecting robust, clinically meaningful, and replicable findings.

Studies will benefit from increased consistency and methodological rigor in both experimental design and equipment. Based on existing data, it cannot be determined to what degree differences in EEG data recording and processing methods (e.g., high versus low impedance EEG systems, electrode density, choice of scalp topography for component extraction) contributed to the diversity of results. Similarly, research on social perception must include appropriate control stimuli. In the research reviewed here, in some cases inference was limited by omission of non-face stimuli, stimuli matched for low-level visual features, and, in studies of emotion, both neutral/natural and emotional faces. Because many studies focused exclusively on social stimuli, it was often impossible to determine whether reported anomalies reflected specific difficulties in social brain circuitry or more general problems with object perception.

Many of these principles have been recognized by researchers and funding agencies, and within-disorder studies incorporating these features are in progress. Though not in the context of electrophysiological markers, the schizophrenia field recognized more than a decade ago with the MATRICS initiative the importance of identifying reliable markers and developing standardized batteries of cognitive tests for use in diagnosis and assessing treatment outcomes (Marder and Fenton 2004). The autism field has more recently begun promoting a similar agenda, with increased focus on the potential utility of electrophysiological biomarkers for capturing heterogeneity and predicting treatment. To this end, the Autism Biomarkers Consortium for Clinical Trials (www.asdbiomarkers.org) is evaluating a battery of EEG and eye-tracking social-communicative biomarkers in a large, rigorously characterized sample of children with ASD, with careful attention to participant characteristics, meticulous control of stimulus, task, and EEG recording parameters across sites, and a longitudinal component to provide information about developmental effects. Likewise, the European Autism Interventions — A Multicentre Study for Developing New Medications (EU-AIMS; Murphy and Spooren 2012), is investigating similar markers, as well as those from additional biomarker domains, such as functional magnetic resonance imaging (fMRI). Although these initiatives are focused on single disorders, their scope and thoroughness are likely to advance understanding of the practical utility of ERP indices of face perception for developing novel interventions and guiding clinical trials.

Finally, the scarcity of studies including multiple diagnostic categories within the same project highlights the urgent need for transdiagnostic work if we are to better understand the specificity versus universality of particular alterations in brain responses to individuals with neurodevelopmental disorders. Thus, moving forward, it is imperative to include both individuals with ASD and SZ – and potentially those with other neurodevelopmental or psychiatric disorders or subthreshold traits – within a single study sample and to measure dimensional traits associated with both disorders across all participants. Studying children or adults at biological risk of ASD or SZ (i.e., siblings, parents, children of affected individuals) to see whether those at risk show altered neural response and/or behavior would also provide important information about intermediate phenotypes and risk states as they relate to brain functioning and clinical profiles. In addition, studies with a longitudinal design and an intervention (targeting either face or emotion processing circuits) may help better parse out causally relevant effects across diagnostic categories.

In summary, both the inconsistencies in findings within ASD and SZ samples and the overlap between findings across ASD and SZ samples make a clear case for the need to consider symptomatology in a dimensional fashion when designing studies and analyzing results in search of biologically meaningful and behaviorally relevant markers of face and emotion processing. Taking a cross-diagnostic, RDoC approach allows for both direct comparison across diagnoses and more detailed sub-grouping or correlations featuring diagnostically cross-cutting, categorically-agnostic phenotypic traits. In addition, this type of study would also allow for clinical characterization to be made along a continuum, rather than imposing an artificial dichotomy (i.e., clinical vs. TD). By taking this approach, future research can address whether and how each categorical disorder (or subset of individuals within a disorder) shows a unique neural signature of altered reception of facial communication, versus whether some alterations in ERP markers of face and emotion processing are reflective of broad clinical impairment in social functioning versus diagnosis or trait-specific pathology. In so doing, research will begin to make more effective headway toward understanding the complexity of distinct versus overlapping etiologies of associated neurodevelopmental disorders, which will in turn position the field to accumulate the precision and replicability of findings needed to develop more targeted treatment approaches.

## Data Availability

All data used for this manuscript are in the included tables.

